# Fewer flights and lower community noise: Noise monitoring at six airports in the United States before, during, and after the COVID-19 pandemic

**DOI:** 10.1101/2025.07.25.25332225

**Authors:** Edmund Seto, Ching-Hsuan Huang

## Abstract

The COVID-19 pandemic resulted in reduced air travel in the United States (US). In the ensuing years after the pandemic, air travel rebounded to near pre-pandemic levels. This provided an opportunity to assess the impact of fewer flights on community noise using monitoring data from six major airports in the US. Flight data for the airports were obtained from the US Department of Transportation (DOT) for the 5-year period before, during, and after the pandemic (2018-2022). Noise levels were assessed at monitoring sites surrounding airports (SFO, LAX, ORD, JFK, LGA, and EWR) in San Francisco, CA; Los Angeles, CA; Chicago, IL; New York City, NY; and Newark, NJ for the same period. Linear models and generalized additive models (GAMs) were used to investigate the changes in Lden and Ldn noise metrics by year and by flight operations. Percentage reduction in flights ranged from 43.9 – 56.2% less in the year of the pandemic (2020) compared to pre-pandemic peak levels. Average noise levels were also found to range from 3.0 – 5.4 dBA lower in 2020 compared to the peak levels before the pandemic. Both flight traffic and noise levels have increased since 2020. Linear regressions and GAMs both indicated lower noise levels at each airport during the pandemic, and associations between numbers of flights and noise levels. Based on linear models, the point estimate effect of air traffic on annual noise level was a 0.8 to 2.7 dBA change per 100,000 annual flights. These findings extend those of previous studies that have also documented noise reductions during the pandemic. This is one of the few studies that evaluated noise reductions in US airport communities in relationship to reduced air traffic across multiple years.

## Introduction

Community noise is an important environmental hazard that is associated with numerous public health issues, including annoyance (1–3), sleep disruption (4,5), cardiovascular health (6), and children’s cognitive skills and performance on standardized tests (7). Aviation contributes to noise exposures in many United States (US) communities, with estimates of over 400,000 people residing in areas with noise levels the U.S. Federal Aviation Administration considers significant (Ldn >65 dBA), a threshold where aircraft noise may create high annoyance for most residents (8). Numerous studies have linked aviation-specific noise to health effects (9), including research suggesting that annoyance from aviation-specific noise is higher than previously quantified (10–12). The Road traffic and Aircraft Noise and children’s Cognition & Health (RANCH) study of children living around London Heathrow, Amsterdam Schiphol, and Madrid Barajas airports advanced our understanding of the association between noise and children’s learning (13,14). Studies of the impact of aviation noise on sleep have found that some noise metrics, such as the maximum sound pressure level or the sound exposure level may be predictive of sleep awakenings, however, variations between field and lab studies suggest that more research on the relationship between aviation noise and sleep is needed (15–17). US research utilizing airport noise contours to assess exposure found increased risk of cardiovascular hospitalization in older adults (18), and a meta-analysis of international studies of noise and hypertension suggests aircraft noise exposure may be associated with increased risk of hypertension (19).

The COVID-19 pandemic dramatically affected air travel in the US. Before the World Health Organization declared COVID-19 a pandemic on March 11, 2020 (20), the US had already implemented procedures to slow the spread of disease, such as routing of flights from affected regions to specific airports in February for screening and potential quarantining (21). By March, the US had implemented additional travel, and some states had begun to implement shutdowns (22–24). By April, most states had reported cases of COVID-19 (24). In addition to the short-term impacts on travel, longer-term shifts in travel behavior post-pandemic include changes in habits and preferences, such as greater options for working from home, commuting less, shopping more online, and taking fewer airplane flights (25).

Various studies have evaluated the impact of the COVID-19 pandemic on changes in community noise levels. Aletta et al. assessed the impact of lockdown measures in London, UK on soundscape recordings, observing reductions in LAeq of 5.4 dB compared to 2019 pre-lockdown levels, with some locations experiencing as much as a 10.7 dB reduction (26).

Asensio et al. measured similar reductions ranging from 4 to 6 dBA for day, evening, and nighttime noise levels during the lockdown in Madrid, Spain (27). Zambon et al. observed reductions of 6 dB, largely due to less transportation-related noise during the lockdown in Milan, Italy (28). Čurović et al. conducted an assessment of the COVID-19 impact on noise levels at the Port of Koper in Slovenia, and observed reductions of 2.2 dB to 5.7 dB compared to long-term averages, and a 23% reduction in people exposed within the 55 dBA noise contour (29).

Walker et al. measured noise in Chelsea, MA, and observed reductions in road and airport-related noise of up to 10 dB during the pandemic (30). Yu et al., used noise data from the Vancouver airport (YVR) and observed reductions of 2.2 dBA during the lockdown, though reductions were less in areas with higher densities of buildings, pavement, and water (31). Amoatey et al. conducted a study in Oman and observed 32% and 35% reductions in LAeq and Lmax, respectively, near the Muscat airport (MCT). None of these previous studies assessed flight reductions or dB reductions at US airports, and most COVID-19 noise studies were focused on short-term (i.e., before and during the pandemic) effects.

The objective of this study was to (1) quantify changes in air traffic and noise in six airport communities over a 5-year period spanning two years before and after the pandemic, and (2) assess the relationship between air traffic and noise in these communities.

## Methods

Flight data for the years 2018 – 2022 were obtained from the Airlines On-Time Performance database which contains both scheduled and actual departure and arrival times reported by certified major US air carriers. The data were downloaded from the US Department of Transportation’s (DOT) Transtats website, and were filtered to exclude cancelled flights and include only those with either an origin or destination that matched the six airports in this study. Actual (rather than scheduled) flight dates and times were used in the analysis. Flights were aggregated to monthly and annual levels for each airport and merged with noise monitoring data.

Noise monitoring data were obtained from each of the six airports (SFO, LAX, ORD, JFK, LGA, and EWR), in the form of online databases or digital (PDF) reports from each airport’s website, from which annual noise metrics were computed. These airports were selected based on the size of their typical flight operations, geographic distribution in the Western, Central, and Eastern regions of the US, and the availability of site-specific noise monitoring data for the 5-year study period. Noise data for JFK, LGA, and EWR were obtained from a single source, The Port Authority of New York and New Jersey, and thus, were considered together in the analyses. California-based SFO and LAX airports provided noise monitoring data using the Community Noise Equivalent Level (CNEL) – a form of the Lden metric used in California, while the other non-California airports provided noise monitoring data using the Ldn metric. While both represent noise metrics in units of dBA, and both are penalized metrics, the Ldn includes a nighttime penalty, while the Lden includes an additional evening time penalty. Because of the differences between Ldn and Lden metrics, statistical models were assessed for each airport individually.

Noise was monitored at 31 sites around SFO airport, 39 sites around LAX, 40 sites around ORD, and 35 sites around JFK, LGA, and EWR. Only SFO provided geographical coordinates for their monitoring sites. For the remaining airports, the locations of monitoring sites were determined by manually geocoding the locations from PDF reports using Google Maps. Maps were created to illustrate the difference between the annual noise level in 2020 and the acoustical average of the pre-pandemic noise levels from 2018 and 2019 for the monitoring sites surrounding each airport.

To assess the association between the outcome of interest (annual noise level in dBA), and the explanatory variable of interest (either categorical year or annual flight traffic), we employed multiple modeling approaches. One set of models used categorical year to assess whether the main pandemic year, 2020 was associated with lower noise. Another set of models used annual flight traffic from the On-Time Performance database as a continuous variable to assess associations with noise level. Multiple linear regression was used to assess the effect of year (or flights), allowing for fixed effects by monitoring site (i.e., categorical site ID).

Additionally, to allow for non-linear effects, a generalized additive model that included a smoothing (spline) term to assess the effect of year (or flights), adjusting for fixed effects by monitoring site, was also considered. Model performance was assessed by Akaike information criterion (AIC) and the R-squared (R^2^).

All analyses were conducted in R version 4.5.0. GAM was performed in R using the mgcv 1.9-3 package. Basemaps were obtained using the maptiles 0.10.0 package and CartoDB.Positron (https://carto.com) tiles, using data from OpenStreetMap (https://www.openstreetmap.org/copyright) downloaded on July 24, 2025.

## Results and Discussion

Flight data confirmed the reduction in flights during the pandemic (Table 1). Depending on the airport, there were between 43.9 – 56.2% fewer flights in 2020 compared to peak annual flights before the pandemic in 2018 - 2019. For all six airports, the number of flights increased between 2020 and 2022, but had not entirely reached the levels pre-pandemic by 2022. Monthly flight traffic data for each airport (Figure 1) illustrate how the flight traffic in January and February of 2020 were similar to the previous years, before a large drop in flights occurred, ultimately reaching the lowest levels in May of 2020 for the 5-year period.

**Table 1.** Annual flights and noise levels by year for each airport.

| Airport | Year | Annual Flights | Ave Noise (dBA)* | SD Noise (dBA)* |
| --- | --- | --- | --- | --- |
| JFK, LGA, EWR | 2018 | 889,218 | 62.9 | 6.6 |
|  | 2019 | 869,810 | 62.2 | 6.9 |
|  | 2020 | 389,919 | 57.9 | 7.6 |
|  | 2021 | 520,523 | 59.0 | 7.4 |
|  | 2022 | 871,535 | 61.0 | 6.9 |
| LAX | 2018 | 442,572 | 64.4 | 4.9 |
|  | 2019 | 439,948 | 64.1 | 4.7 |
|  | 2020 | 248,365 | 61.4 | 4.8 |
|  | 2021 | 348,553 | 63.7 | 3.8 |
|  | 2022 | 382,319 | 63.8 | 3.7 |
| ORD | 2018 | 665,894 | 59.8 | 5.7 |
|  | 2019 | 679,175 | 59.8 | 5.9 |
|  | 2020 | 379,237 | 56.3 | 5.9 |
|  | 2021 | 481,881 | 57.8 | 6.5 |
|  | 2022 | 521,158 | 58.2 | 6.4 |
| SFO | 2018 | 351,664 | 65.8 | 8.7 |
|  | 2019 | 341,884 | 65.0 | 6.5 |
|  | 2020 | 178,817 | 60.4 | 3.7 |
|  | 2021 | 197,572 | 61.8 | 3.6 |
|  | 2022 | 259,913 | 61.7 | 4.2 |

\* Lden are reported for SFO and LAX, Ldn are reported for ORD and JFK, LGA, and EWR. Mean and SD are computed across the annual Lden (or Ldn) for monitoring sites at each airport.

**Figure 1.**
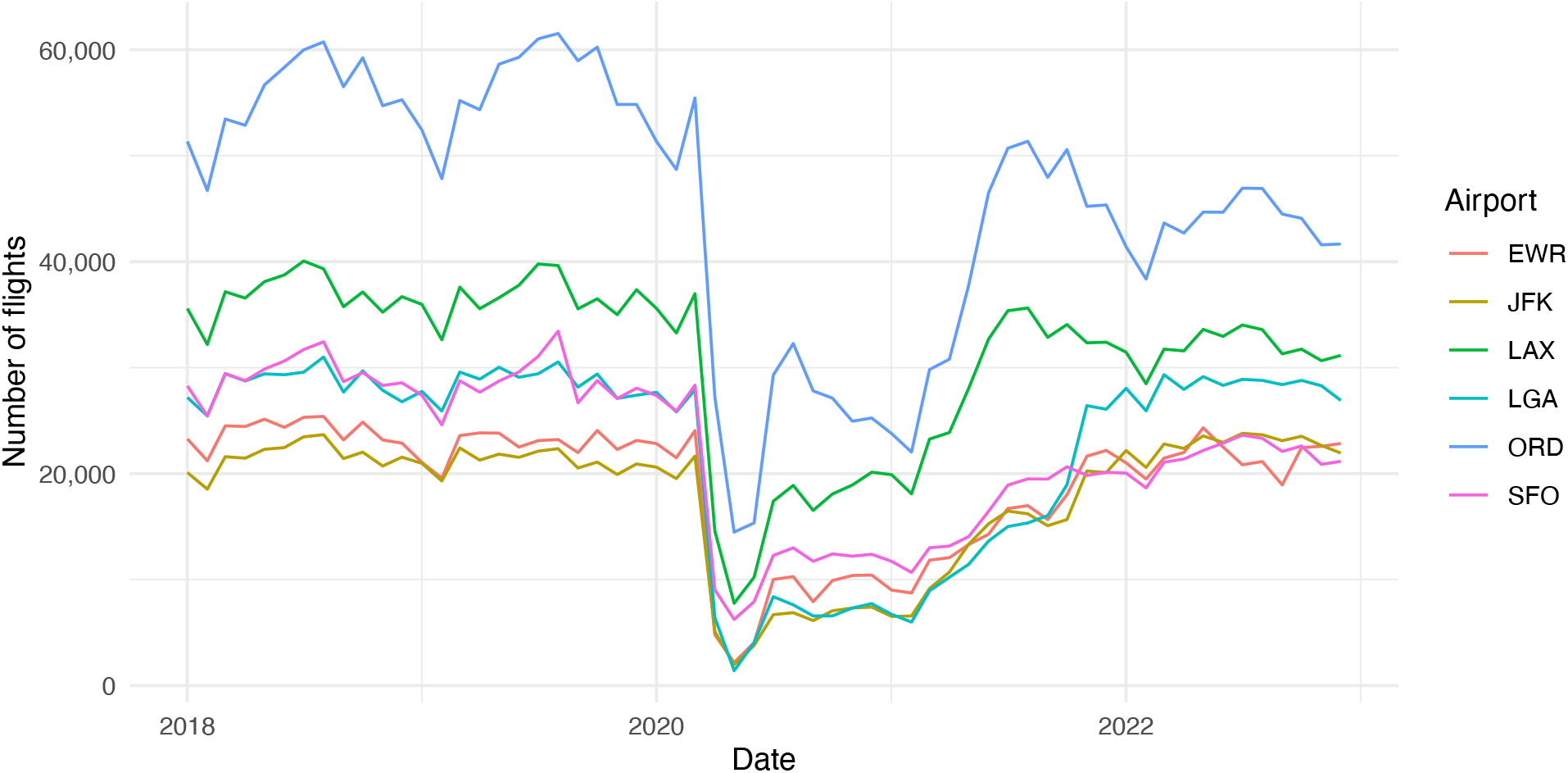
Monthly flight operations reported by major commercial carriers to the Federal Aviation Administration at six airports for years 2018 to 2022.

The noise levels at all six airports were also lowest during the pandemic year (Table 1), ranging from 57.9 dBA (Ldn) at the JFK, LGA, and EWR airports to 61.4 dBA (Lden) at the LAX airport. These levels were between 3.0 – 5.4 dBA lower than noise levels observed in the two pre-pandemic years. Similar to the flight traffic data, annual noise levels have increased at each airport from 2020 to 2022, not quite reaching the levels before the pandemic.

The drop in noise during the pandemic was also observed in the results of the GAM with the smoothing spline term for categorical year (Tables S1 – S4 and Figure 2). Plots of the smoothing spline term by year (Figure 2) illustrate the drop in noise for the middle to late 2020 period, gradually recovering in subsequent years. Linear regression models that included categorical year (Tables S5 – S8) also indicated significant associations between the year 2020 and lower noise levels at each airport.

**Figure 2.**
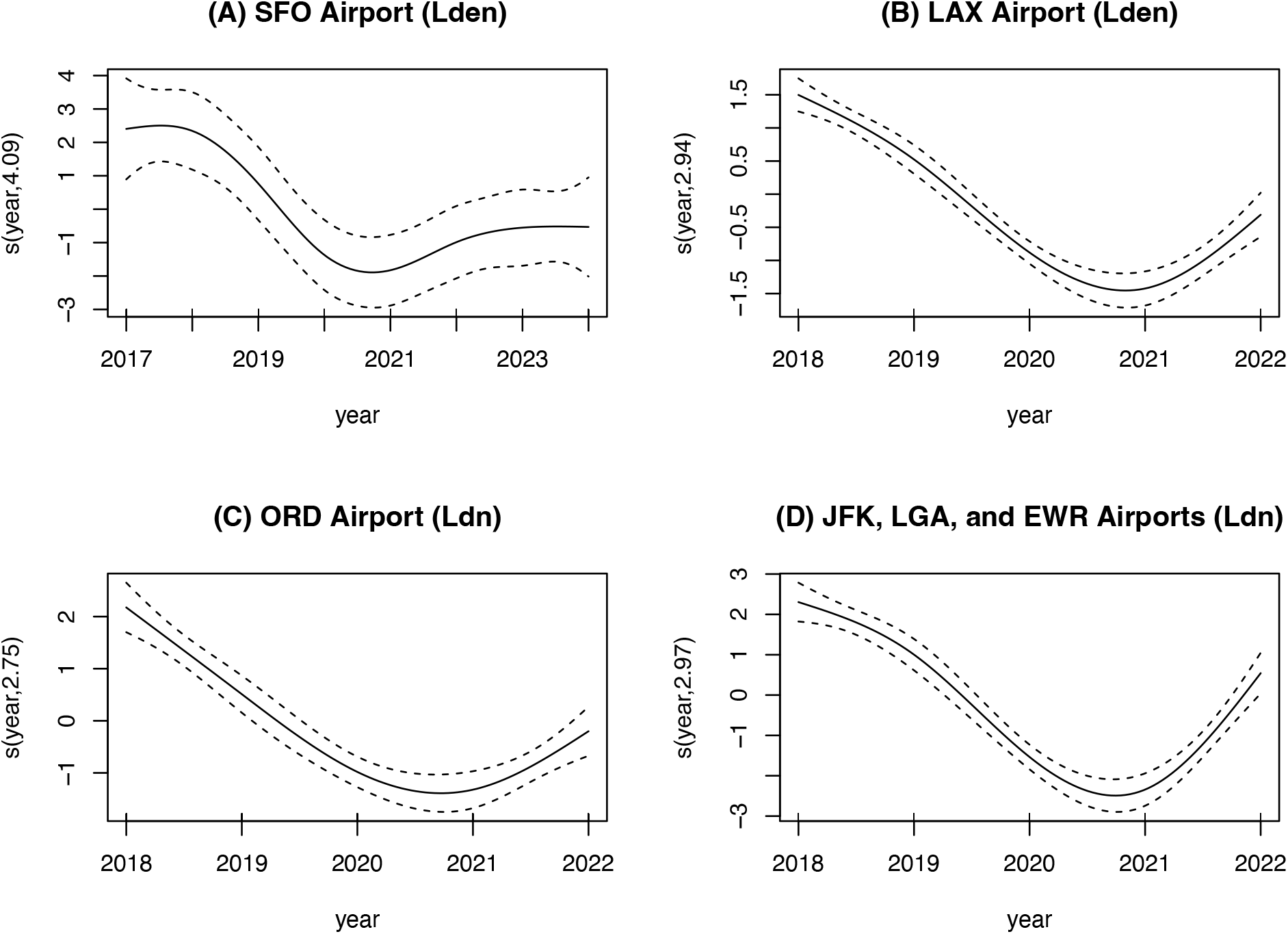
Smoothing component illustrating the effect of year for the Generalized Additive Model fit to Lden and Ldn noise monitoring data for airports for years before, during, and after the COVID-19 pandemic.

Although the most pronounced reduction in noise and air traffic occurred during 2020, there were variations in noise and traffic throughout the 5-year period. The results of the GAM with a smoothing spline term for flight traffic indicate a strong relationship between numbers of flights and noise at each airport (Tables S9 – S12 and Figure 3). The GAM smoothing component for flight traffic illustrates an approximately linear effect of increasing flights on increased noise (Figure 3). The coefficients of the corresponding linear regression models (the “per 1,000 flights”, nper1000 variable in Tables S13 – S16) suggests an increase of 0.008 to0.027 dBA for every 1,000 additional flights (or rescaled: 0.8 to 2.7 dBA increase per 100,000 additional flights).

**Figure 3.**
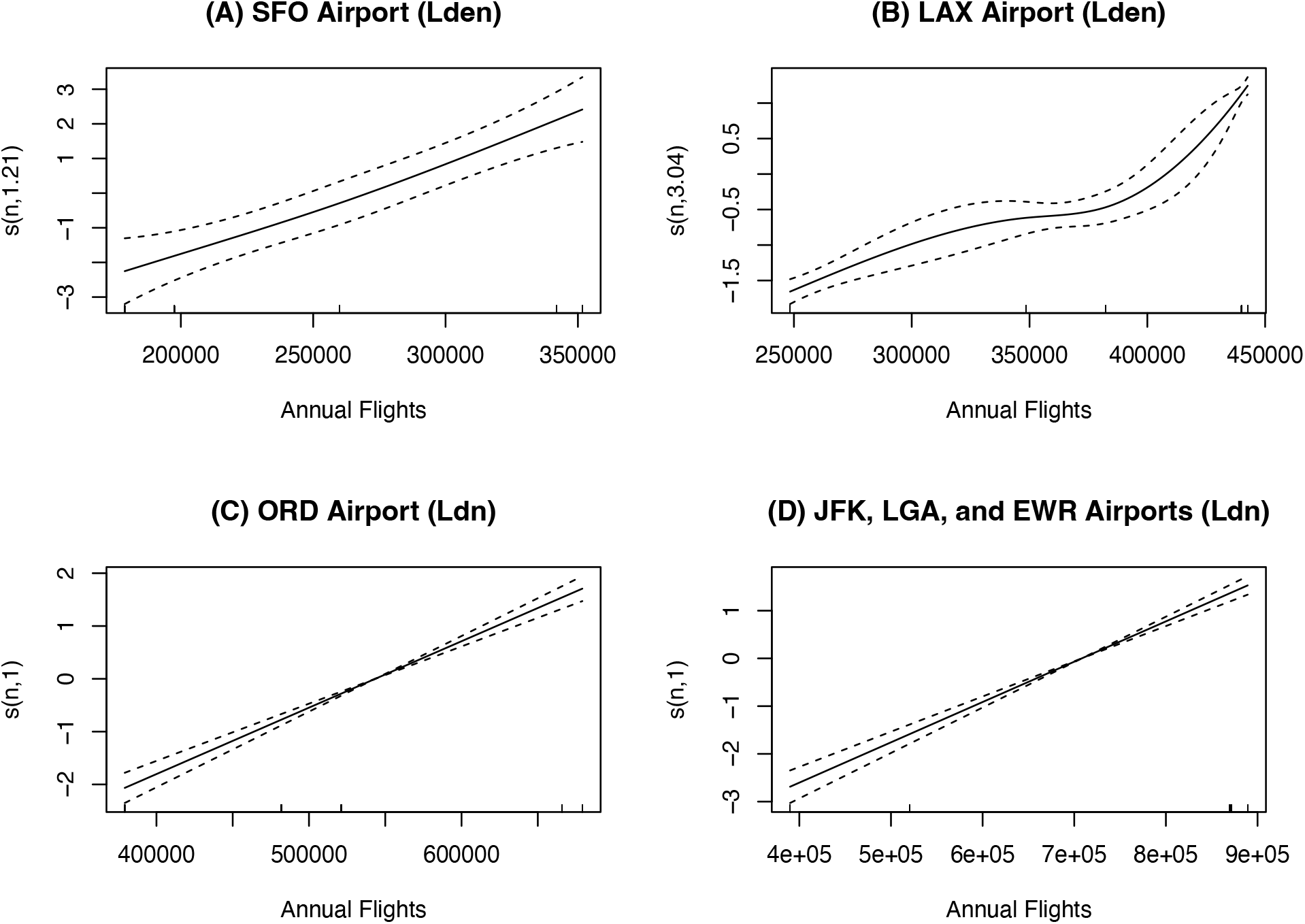
Smoothing component illustrating the effect of annual flights for the Generalized Additive Model fit to Lden and Ldn noise monitoring data for airports for years 2018-2022.

Comparisons of the AIC and R^2^ performance statistics for the various airport noise models (Table S17) suggest that both GAM and linear models performed well in most cases with R^2^ ≥ 0.94, except for the noise model for SFO, which only resulted in R^2^ between 0.41 and 0.57 (see later discussion of the map of SFO noise data, and restricted model sensitivity analysis). The AICs were slightly better (lower) for some of the GAM models compared to the corresponding linear regression version. But generally, both model forms captured the year and traffic effects on noise. This was likely due to the clear annual differences in traffic and noise by year across airports, as well as the near linear smoothing spline for traffic.

The spatial patterns of noise reductions observed during the main pandemic year 2020 relative to peak levels in previous years provide useful insights into the overall regional noise reductions and peculiarities of certain noise monitoring sites (Figure 4). Each airport location is depicted as a red-colored triangle on the map panels. Blue circles indicate the location of noise monitors, with their size depicting the dB reduction in 2020 compared to peak levels in 2018-2019. Notably, there were reductions in noise levels at all sites, and at all the airports, including sites closest to the airport as well as those farther away. This suggests that community noise level reductions during the pandemic may also be attributed to reductions in other noise sources, including other forms of transportation (e.g., road and rail) and social events and gatherings. This confounding may be difficult to disentangle; although the statistical models suggest significant associations between reduced air traffic and lower community noise, region-wide changes in transportation and social activities likely occurred at the same time during the pandemic. As a sensitivity analysis to assess the effect of air traffic on community noise in the absence of the pandemic, we excluded data for the pandemic year 2020 and re-ran the linear models (Tables S18 – S21). After exclusion, we still observed significant positive associations between air traffic and noise with similar effect sizes.

**Figure 4.**
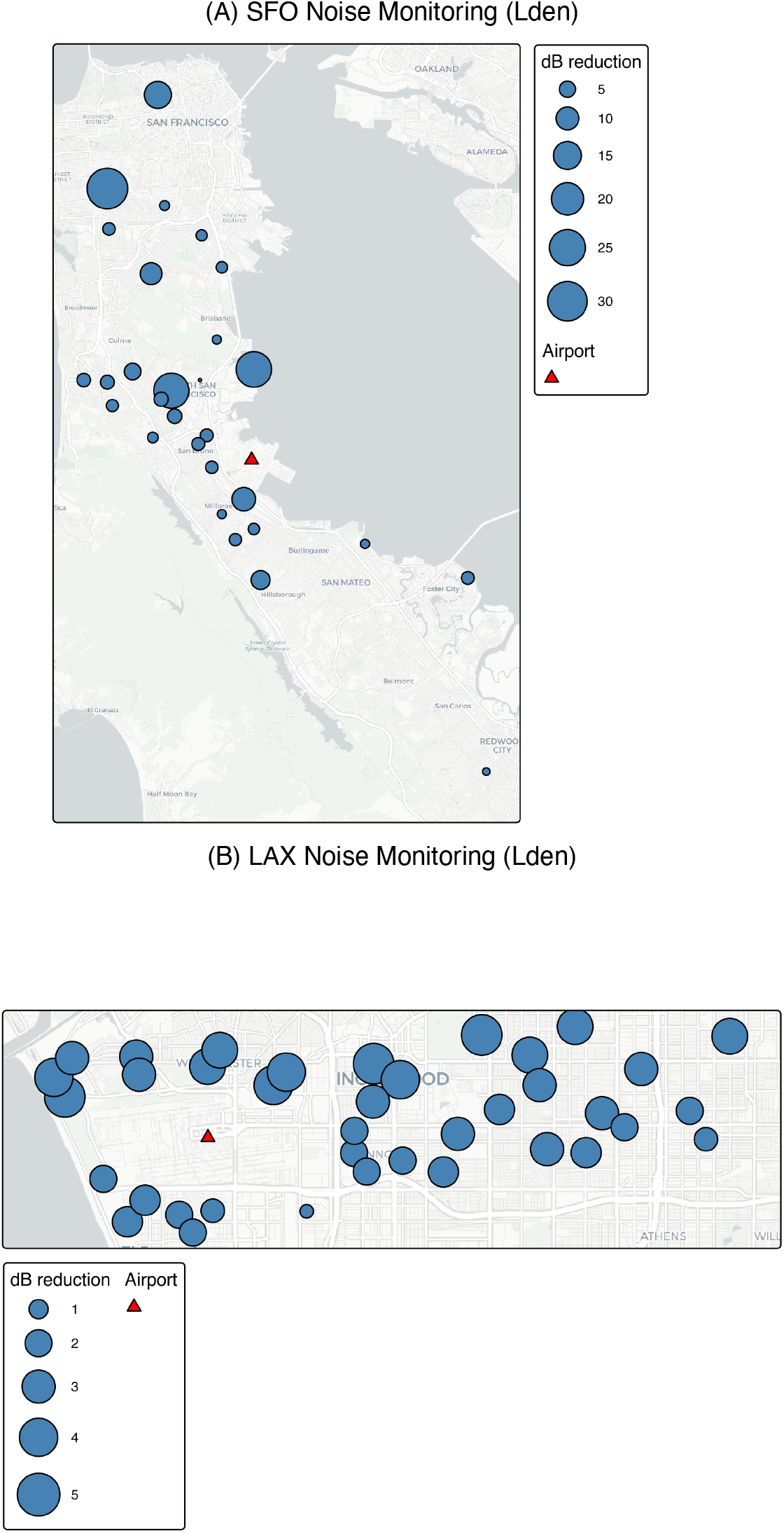
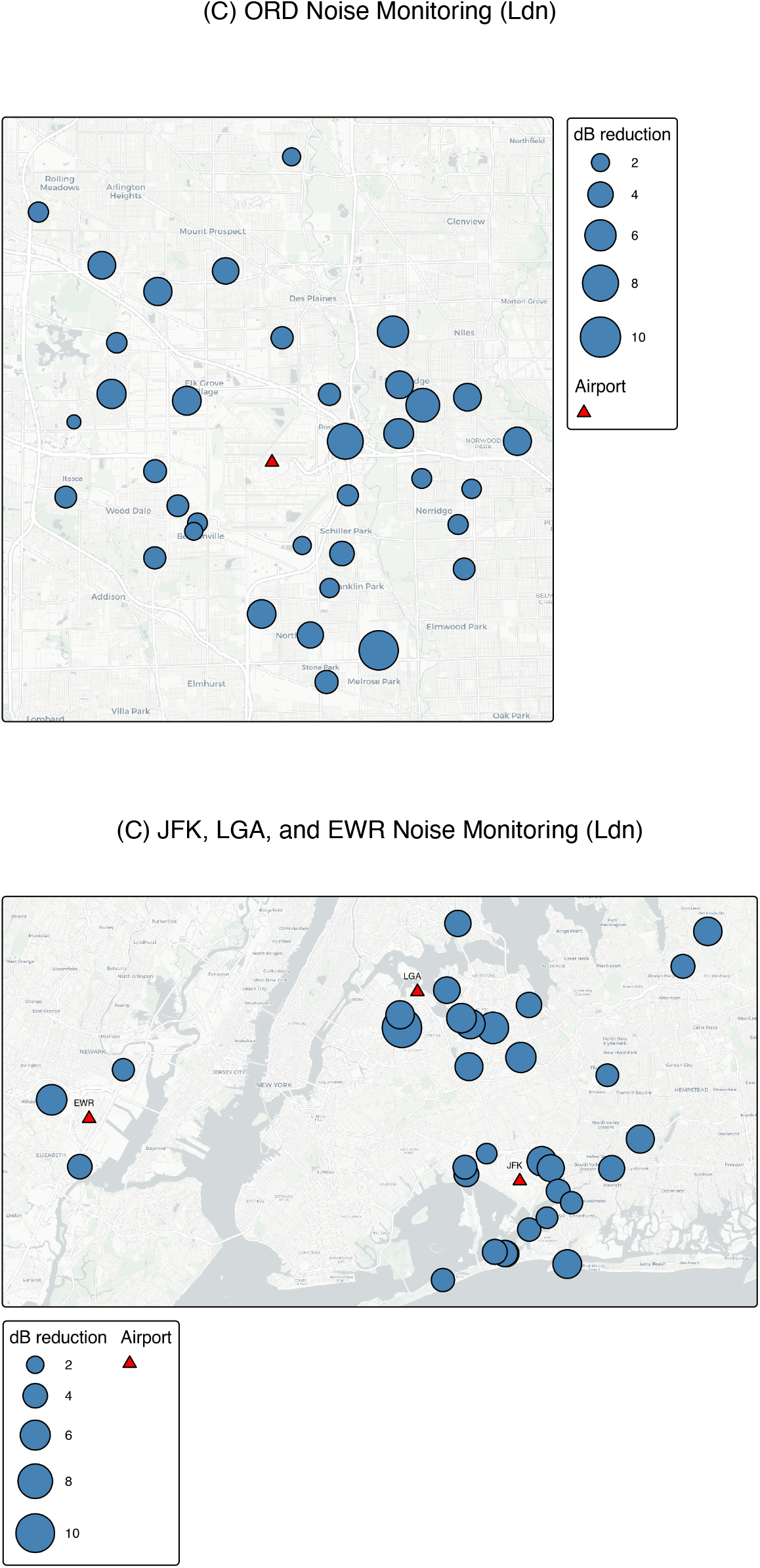
Maps of reductions in Lden and Ldn noise levels (dBA) in year 2020 versus 2018-2019 noise levels for monitoring sites at each airport. Basemap from CARTO, using data from OpenStreetMaps.

The map for the San Francisco area (Figure 4, panel (A)) illustrates some site-specific peculiarities that are not as prevalent in the other airport regions. A few monitoring sites for SFO had relatively large dB reductions. These sites were not particularly close to the airport, with one of the sites located in the Golden Gate Heights District in the city of San Francisco, one near Oyster Point Marina/Park, and another near a major arterial street (CA State Route 82) in South San Francisco. These sites had relatively high pre-pandemic noise levels exceeding 80 dBA, presumably due to non-aviation noise sources. Excluding these three sites and year 2020 in a sensitivity analysis with a linear model, resulted in an improved R^2^ of 0.77, yet a significant positive association between air traffic and noise level for the remaining sites was still observed (Table S22).

The limitations of this study include the consideration of only six airports, the limited flight data availability, the limited ability to isolate air traffic from other sources of noise, the lack of consideration of human exposures or population health impacts, and limited availability and inconsistencies in noise metrics reported by airports. First, while flight data were available for the six airports in this study, and many other airports in the US from the On-Time Performance database, these data serve as an indicator for flight traffic rather than an accurate count of all flight operations, as the data do not include aircraft that are not required to be reported in the On-Time Performance to the US DOT, such as freight, military, and non-commercial aviation.

As such, the On-Time Performance data is an undercount of actual flights at airports. Second, unfortunately, there is no single consolidated and homogeneous noise monitoring and reporting system for the US. Therefore, noise data for this study were obtained from each monitoring agency and individually processed. The lack of consistent noise reporting also contributed to discrepancies in noise metrics, with airports reporting Lden or Ldn, but not both. Other major US airports, such as those in Dallas, TX, Denver, CO and Atlanta, GA have Noise and Operations Monitoring Systems (NOMS), which allow public access to real-time and historical noise monitoring and flight tracking data for filing noise complaints. However, their NOMS do not provide a way to download historical noise data for research. Moreover, summary noise metrics, such as the Lden and Lden do not allow for listening studies and soundscape analyses that would help to better identify and quantify reductions in particular noise sources, which would be possible with historical sound recordings. There would be opportunities for more research if routine noise monitoring data were more systematically collected, maintained, and shared by airport agencies. Finally, while this study did not consider changes in population noise exposures and health impacts, existing aviation-specific noise exposure response functions (ERFs) may be useful for quantifying the health impacts associated with the reductions in noise found in this study. However, as these reductions occurred during an unusual global pandemic, it is unclear whether previously estimated ERFs would be appropriate to use for a period when noise perceptions may have shifted, and various competing health risks were prevalent.

## Conclusion

This is one of the few studies that has considered the relationship between air traffic and noise at multiple airport communities before, during, and after the COVID-19 pandemic in the US. Key findings include concomitant reductions in air traffic and noise during the period of the pandemic, and the quantification of a significant positive linear relationship between air traffic and community noise in the six airport communities.

## Supporting information

GAM results for year

GAM results for flight traffic

LM results for year

LM results for restricted model without 2020 flight traffic

LM results for flight traffic

LM results for restricted model for SFO

GAM and LM model comparisons

## Data Availability

All data produced are available online at:
https://data.sfgov.org/Transportation/Aircraft-Noise-Climates/qxw2-ncq3/about_data
https://www.lawa.org/lawa-environment/noise-management/lawa-noise-management-lax/california-state-airport-noise-standards-quarterly-reports-and-contour-maps
https://www.flychicago.com/community/ORDnoise/ANMS/Pages/ANMSreports.aspx
https://aircraftnoise.panynj.gov/reports/

https://www.lawa.org/lawa-environment/noise-management/lawa-noise-management-lax/california-state-airport-noise-standards-quarterly-reports-and-contour-maps

https://www.flychicago.com/community/ORDnoise/ANMS/Pages/ANMSreports.aspx

https://aircraftnoise.panynj.gov/reports/

## Acknowledgements

We thank the US DOT for making On-Time Performance database available via the Transtats website. We thank SFO, LAX, ORD and The Port Authority of New York and New Jersey for making noise data available on their websites.

