## Supplementary material for "Fewer flights and lower community noise: Noise monitoring at six airports in the United States before, during, and after the COVID-19 pandemic": GAM results for year

**Table S1.** Generalized Additive Model Summary for SFO Lden (dBA) noise levels with a smoothing spline on year.

| Component | Term | Estimate | Std Error | t-value | p-value | |
|---|---|---|---|---|---|---|
| A. parametric coefficients | (Intercept) | 73.144 | 1.596 | 45.824 | 0.0000 | *** |
| | factor(location_id)2 | -8.511 | 2.444 | -3.483 | 0.0006 | *** |
| | factor(location_id)3 | -10.050 | 2.257 | -4.452 | 0.0000 | *** |
| | factor(location_id)4 | -5.650 | 2.257 | -2.503 | 0.0131 | * |
| | factor(location_id)5 | -6.100 | 2.257 | -2.702 | 0.0075 | ** |
| | factor(location_id)6 | -8.600 | 2.257 | -3.810 | 0.0002 | *** |
| | factor(location_id)7 | -12.762 | 2.257 | -5.654 | 0.0000 | *** |
| | factor(location_id)8 | -4.362 | 2.257 | -1.933 | 0.0547 | . |
| | factor(location_id)9 | -14.362 | 2.257 | -6.363 | 0.0000 | *** |
| | factor(location_id)10 | -14.225 | 2.257 | -6.302 | 0.0000 | *** |
| | factor(location_id)11 | -14.550 | 2.257 | -6.446 | 0.0000 | *** |
| | factor(location_id)12 | -9.737 | 2.257 | -4.314 | 0.0000 | *** |
| | factor(location_id)13 | -14.862 | 2.257 | -6.584 | 0.0000 | *** |
| | factor(location_id)14 | -6.887 | 2.257 | -3.051 | 0.0026 | ** |
| | factor(location_id)15 | -7.337 | 2.257 | -3.251 | 0.0014 | ** |
| | factor(location_id)16 | -8.212 | 2.257 | -3.638 | 0.0004 | *** |
| | factor(location_id)17 | -11.212 | 2.257 | -4.967 | 0.0000 | *** |
| | factor(location_id)18 | -7.850 | 2.257 | -3.478 | 0.0006 | *** |
| | factor(location_id)19 | -11.112 | 2.257 | -4.923 | 0.0000 | *** |
| | factor(location_id)20 | -10.525 | 2.257 | -4.663 | 0.0000 | *** |
| | factor(location_id)21 | -12.337 | 2.257 | -5.466 | 0.0000 | *** |
| | factor(location_id)22 | -8.175 | 2.257 | -3.622 | 0.0004 | *** |
| | factor(location_id)23 | -8.237 | 2.257 | -3.649 | 0.0003 | *** |
| | factor(location_id)24 | -9.112 | 2.257 | -4.037 | 0.0001 | *** |
| | factor(location_id)25 | -15.987 | 2.257 | -7.082 | 0.0000 | *** |
| | factor(location_id)26 | -8.025 | 2.257 | -3.555 | 0.0005 | *** |
| | factor(location_id)27 | -11.250 | 2.257 | -4.984 | 0.0000 | *** |
| | factor(location_id)28 | -10.530 | 2.780 | -3.788 | 0.0002 | *** |
| | factor(location_id)29 | -13.887 | 2.257 | -6.152 | 0.0000 | *** |
| | factor(location_id)281 | -16.883 | 3.076 | -5.488 | 0.0000 | *** |
| | factor(location_id)282 | -18.921 | 3.079 | -6.145 | 0.0000 | *** |
| **Component** | **Term** | **edf** | **Ref. df** | **F-value** | **p-value** | |

| Component | Term | Estimate | Std Error | t-value | p-value | |
|---|---|---|---|---|---|---|
| B. smooth terms | s(year) | 4.089 | 4.863 | 5.511 | 0.0001 | *** |

Signif. codes: 0 <= '***' < 0.001 < '**' < 0.01 < '*' < 0.05

Adjusted R-squared: 0.407, Deviance explained 0.494

GCV : 24.014, Scale est: 20.382, N: 232

**Table S2.** Generalized Additive Model Summary for LAX Lden (dBA) noise levels with a smoothing spline on year.

| Component | Term | Estimate | Std Error | t-value | p-value | |
|---|---|---|---|---|---|---|
| A. parametric coefficients | (Intercept) | 77.953 | 0.513 | 151.924 | 0.0000 | *** |
| | factor(SiteID)ATH1 | -17.667 | 0.723 | -24.444 | 0.0000 | *** |
| | factor(SiteID)ATH2 | -12.034 | 0.649 | -18.536 | 0.0000 | *** |
| | factor(SiteID)DEL1 | -20.234 | 0.649 | -31.167 | 0.0000 | *** |
| | factor(SiteID)ESG1 | -13.434 | 0.649 | -20.693 | 0.0000 | *** |
| | factor(SiteID)ESG2 | -10.434 | 0.649 | -16.072 | 0.0000 | *** |
| | factor(SiteID)ESG3 | -15.834 | 0.649 | -24.390 | 0.0000 | *** |
| | factor(SiteID)ESG4 | -19.000 | 0.723 | -26.289 | 0.0000 | *** |
| | factor(SiteID)ESG5 | -17.234 | 0.649 | -26.546 | 0.0000 | *** |
| | factor(SiteID)ING1 | -17.234 | 0.649 | -26.546 | 0.0000 | *** |
| | factor(SiteID)ING2 | -12.234 | 0.649 | -18.845 | 0.0000 | *** |
| | factor(SiteID)ING3 | -11.634 | 0.649 | -17.920 | 0.0000 | *** |
| | factor(SiteID)ING5 | -18.667 | 0.723 | -25.827 | 0.0000 | *** |
| | factor(SiteID)ING6 | -8.434 | 0.649 | -12.991 | 0.0000 | *** |
| | factor(SiteID)ING7 | -21.333 | 0.723 | -29.517 | 0.0000 | *** |
| | factor(SiteID)ING8 | -16.634 | 0.649 | -25.622 | 0.0000 | *** |
| | factor(SiteID)LNX1 | -3.834 | 0.649 | -5.906 | 0.0000 | *** |
| | factor(SiteID)LNX2 | -15.434 | 0.649 | -23.773 | 0.0000 | *** |
| | factor(SiteID)LNX3 | -15.634 | 0.649 | -24.082 | 0.0000 | *** |
| | factor(SiteID)LNX4 | -12.834 | 0.649 | -19.769 | 0.0000 | *** |
| | factor(SiteID)PDR1 | -10.234 | 0.649 | -15.764 | 0.0000 | *** |
| | factor(SiteID)PDR2 | -15.834 | 0.649 | -24.390 | 0.0000 | *** |
| | factor(SiteID)PDR3 | -20.333 | 0.723 | -28.134 | 0.0000 | *** |
| | factor(SiteID)SLA1 | -13.634 | 0.649 | -21.001 | 0.0000 | *** |
| | factor(SiteID)SLA2 | -19.333 | 0.723 | -26.750 | 0.0000 | *** |
| | factor(SiteID)SLA3 | -16.234 | 0.649 | -25.006 | 0.0000 | *** |
| | factor(SiteID)SLA4 | -19.000 | 0.723 | -26.289 | 0.0000 | *** |
| | factor(SiteID)SLA5 | -13.834 | 0.649 | -21.309 | 0.0000 | *** |
| | factor(SiteID)SLA6 | -16.333 | 0.723 | -22.599 | 0.0000 | *** |
| | factor(SiteID)SLA7 | -13.634 | 0.649 | -21.001 | 0.0000 | *** |
| | factor(SiteID)SLA8 | -17.333 | 0.723 | -23.983 | 0.0000 | *** |
| | factor(SiteID)SLA9 | -17.000 | 0.723 | -23.521 | 0.0000 | *** |

| Component | Term | Estimate | Std Error | t-value | p-value | |
|---|---|---|---|---|---|---|
| | factor(SiteID)WCH1 | -21.333 | 0.723 | -29.517 | 0.0000 | *** |
| | factor(SiteID)WCH2 | -16.034 | 0.649 | -24.698 | 0.0000 | *** |
| | factor(SiteID)WCH3 | -16.834 | 0.649 | -25.930 | 0.0000 | *** |
| | factor(SiteID)WCH4 | -20.000 | 0.723 | -27.672 | 0.0000 | *** |
| | factor(SiteID)WCH5 | -4.834 | 0.649 | -7.446 | 0.0000 | *** |
| | factor(SiteID)WCH6 | -15.434 | 0.649 | -23.773 | 0.0000 | *** |

| Component | Term | edf | Ref. df | F-value | p-value | |
|---|---|---|---|---|---|---|
| B. smooth terms | s(year) | 2.940 | 2.997 | 74.090 | 0.0000 | *** |

Signif. codes: 0 <= '***' < 0.001 < '**' < 0.01 < '*' < 0.05

Adjusted R-squared: 0.963, Deviance explained 0.972

GCV : 1.044, Scale est: 0.784, N: 164

**Table S3.** Generalized Additive Model Summary for ORD Ldn (dBA) noise levels with a smoothing spline on year.

| Component | Term | Estimate | Std Error | t-value | p-value | |
|---|---|---|---|---|---|---|
| A. parametric coefficients | (Intercept) | 52.184 | 0.692 | 75.419 | 0.0000 | *** |
| | factor(SiteID)10 | -4.020 | 0.979 | -4.108 | 0.0001 | *** |
| | factor(SiteID)11 | 0.760 | 0.979 | 0.777 | 0.4387 | |
| | factor(SiteID)12 | 5.620 | 0.979 | 5.743 | 0.0000 | *** |
| | factor(SiteID)13 | 10.960 | 0.979 | 11.201 | 0.0000 | *** |
| | factor(SiteID)14 | 2.600 | 0.979 | 2.657 | 0.0088 | ** |
| | factor(SiteID)15 | 0.100 | 0.979 | 0.102 | 0.9188 | |
| | factor(SiteID)16 | 11.460 | 0.979 | 11.712 | 0.0000 | *** |
| | factor(SiteID)17 | 9.820 | 0.979 | 10.036 | 0.0000 | *** |
| | factor(SiteID)18 | 6.065 | 1.039 | 5.834 | 0.0000 | *** |
| | factor(SiteID)19 | 2.580 | 0.979 | 2.637 | 0.0094 | ** |
| | factor(SiteID)2 | 2.240 | 0.979 | 2.289 | 0.0236 | * |
| | factor(SiteID)20 | 0.880 | 0.979 | 0.899 | 0.3701 | |
| | factor(SiteID)21 | -2.404 | 1.040 | -2.313 | 0.0223 | * |
| | factor(SiteID)22 | 12.440 | 0.979 | 12.713 | 0.0000 | *** |
| | factor(SiteID)23 | 7.840 | 0.979 | 8.012 | 0.0000 | *** |
| | factor(SiteID)24 | 1.820 | 0.979 | 1.860 | 0.0651 | . |
| | factor(SiteID)25 | 9.980 | 0.979 | 10.199 | 0.0000 | *** |
| | factor(SiteID)26 | 4.680 | 0.979 | 4.783 | 0.0000 | *** |
| | factor(SiteID)27 | 16.000 | 0.979 | 16.352 | 0.0000 | *** |
| | factor(SiteID)28 | 20.880 | 0.979 | 21.339 | 0.0000 | *** |
| | factor(SiteID)29 | 14.760 | 0.979 | 15.084 | 0.0000 | *** |
| | factor(SiteID)3 | 11.380 | 0.979 | 11.630 | 0.0000 | *** |
| | factor(SiteID)30 | 3.596 | 1.040 | 3.459 | 0.0007 | *** |
| | factor(SiteID)31 | 18.590 | 1.039 | 17.883 | 0.0000 | *** |
| | factor(SiteID)32 | 11.500 | 0.979 | 11.753 | 0.0000 | *** |
| | factor(SiteID)33 | 9.020 | 0.979 | 9.218 | 0.0000 | *** |
| | factor(SiteID)34 | 5.240 | 0.979 | 5.355 | 0.0000 | *** |
| | factor(SiteID)35 | 3.740 | 0.979 | 3.822 | 0.0002 | *** |
| | factor(SiteID)37 | -3.480 | 0.979 | -3.556 | 0.0005 | *** |
| | factor(SiteID)39 | 8.117 | 1.711 | 4.743 | 0.0000 | *** |
| | factor(SiteID)4 | 3.420 | 0.979 | 3.495 | 0.0006 | *** |

| Component | Term | Estimate | Std Error | t-value | p-value | |
|---|---|---|---|---|---|---|
| | factor(SiteID)40 | 2.117 | 1.711 | 1.237 | 0.2182 | |
| | factor(SiteID)42 | 4.860 | 0.979 | 4.967 | 0.0000 | *** |
| | factor(SiteID)44 | 11.365 | 1.039 | 10.933 | 0.0000 | *** |
| | factor(SiteID)45 | -1.183 | 1.711 | -0.691 | 0.4906 | |
| | factor(SiteID)46 | 3.917 | 1.711 | 2.289 | 0.0236 | * |
| | factor(SiteID)5 | 5.696 | 1.040 | 5.479 | 0.0000 | *** |
| | factor(SiteID)7 | -0.829 | 1.040 | -0.798 | 0.4265 | |
| | factor(SiteID)8 | 5.000 | 0.979 | 5.110 | 0.0000 | *** |
| **Component** | **Term** | **edf** | **Ref. df** | **F-value** | **p-value** | |
| B. smooth terms | s(year) | 2.748 | 2.951 | 39.769 | 0.0000 | *** |

Signif. codes: 0 <= '***' < 0.001 < '**' < 0.01 < '*' < 0.05

Adjusted R-squared: 0.938, Deviance explained 0.952

GCV : 3.156, Scale est: 2.394, N: 177

**Table S4.** Generalized Additive Model Summary for JFK, LGA, and EWR Ldn (dBA) noise levels with a smoothing spline on year.

| Component | Term | Estimate | Std Error | t-value | p-value | |
|---|---|---|---|---|---|---|
| A. parametric coefficients | (Intercept) | 70.166 | 0.680 | 103.117 | 0.0000 | *** |
| | factor(SiteID)2 | 0.080 | 0.962 | 0.083 | 0.9339 | |
| | factor(SiteID)3 | -25.100 | 0.962 | -26.084 | 0.0000 | *** |
| | factor(SiteID)4 | -5.440 | 0.962 | -5.653 | 0.0000 | *** |
| | factor(SiteID)5 | 0.160 | 0.962 | 0.166 | 0.8682 | |
| | factor(SiteID)6 | -1.800 | 0.962 | -1.871 | 0.0638 | . |
| | factor(SiteID)7 | -8.260 | 0.962 | -8.584 | 0.0000 | *** |
| | factor(SiteID)8 | -5.160 | 0.962 | -5.362 | 0.0000 | *** |
| | factor(SiteID)9 | -3.700 | 0.962 | -3.845 | 0.0002 | *** |
| | factor(SiteID)11 | -1.980 | 0.962 | -2.058 | 0.0418 | * |
| | factor(SiteID)28 | -3.240 | 0.962 | -3.367 | 0.0010 | ** |
| | factor(SiteID)52 | -15.020 | 0.962 | -15.609 | 0.0000 | *** |
| | factor(SiteID)54 | -15.540 | 0.962 | -16.149 | 0.0000 | *** |
| | factor(SiteID)55 | -9.220 | 0.962 | -9.581 | 0.0000 | *** |
| | factor(SiteID)56 | -3.200 | 0.962 | -3.325 | 0.0012 | ** |
| | factor(SiteID)57 | -4.273 | 1.023 | -4.179 | 0.0001 | *** |
| | factor(SiteID)58 | -7.080 | 0.962 | -7.358 | 0.0000 | *** |
| | factor(SiteID)59 | -7.100 | 0.962 | -7.378 | 0.0000 | *** |
| | factor(SiteID)62 | -9.920 | 0.962 | -10.309 | 0.0000 | *** |
| | factor(SiteID)66 | -14.080 | 0.962 | -14.632 | 0.0000 | *** |
| | factor(SiteID)67 | -10.760 | 0.962 | -11.182 | 0.0000 | *** |
| | factor(SiteID)68 | -9.769 | 1.281 | -7.627 | 0.0000 | *** |
| | factor(SiteID)69 | -19.300 | 0.962 | -20.056 | 0.0000 | *** |
| | factor(SiteID)71 | -10.300 | 0.962 | -10.704 | 0.0000 | *** |
| | factor(SiteID)72 | -16.900 | 0.962 | -17.562 | 0.0000 | *** |
| | factor(SiteID)74 | -2.860 | 0.962 | -2.972 | 0.0036 | ** |
| | factor(SiteID)76 | -16.269 | 1.281 | -12.701 | 0.0000 | *** |
| | factor(SiteID)77 | -16.920 | 0.962 | -17.583 | 0.0000 | *** |
| | factor(SiteID)78 | -20.700 | 0.962 | -21.511 | 0.0000 | *** |
| | factor(SiteID)79 | -12.348 | 1.023 | -12.076 | 0.0000 | *** |
| | factor(SiteID)80 | -17.720 | 0.962 | -18.415 | 0.0000 | *** |
| | factor(SiteID)82 | -3.200 | 0.962 | -3.325 | 0.0012 | ** |

| Component | Term | Estimate | Std Error | t-value | p-value | |
|---|---|---|---|---|---|---|
| | factor(SiteID)84 | -16.132 | 1.022 | -15.779 | 0.0000 | *** |
| | factor(SiteID)85 | -19.240 | 1.115 | -17.251 | 0.0000 | *** |
| | factor(SiteID)86 | -14.408 | 1.685 | -8.550 | 0.0000 | *** |
| **Component** | **Term** | **edf** | **Ref. df** | **F-value** | **p-value** | |
| B. smooth terms | s(year) | 2.966 | 2.999 | 65.769 | 0.0000 | *** |

Signif. codes: 0 <= '***' < 0.001 < '**' < 0.01 < '*' < 0.05

Adjusted R-squared: 0.956, Deviance explained 0.966

GCV : 3.035, Scale est: 2.315, N: 160
