## Supplementary material for "Fewer flights and lower community noise: Noise monitoring at six airports in the United States before, during, and after the COVID-19 pandemic": GAM results for flight traffic

**Table S9.** Generalized Additive Model Summary for SFO Lden (dBA) noise levels with a smoothing spline on flight traffic (denoted as variable n).

| Component | Term | Estimate | Std Error | t-value | p-value | |
|---|---|---|---|---|---|---|
| A. parametric coefficients | (Intercept) | 72.666 | 1.975 | 36.787 | 0.0000 | *** |
| | factor(location_id)2 | -8.455 | 2.965 | -2.851 | 0.0052 | ** |
| | factor(location_id)3 | -9.640 | 2.794 | -3.451 | 0.0008 | *** |
| | factor(location_id)4 | -5.720 | 2.794 | -2.048 | 0.0429 | * |
| | factor(location_id)5 | -6.000 | 2.794 | -2.148 | 0.0338 | * |
| | factor(location_id)6 | -8.600 | 2.794 | -3.079 | 0.0026 | ** |
| | factor(location_id)7 | -11.900 | 2.794 | -4.260 | 0.0000 | *** |
| | factor(location_id)8 | -2.900 | 2.794 | -1.038 | 0.3014 | |
| | factor(location_id)9 | -13.960 | 2.794 | -4.997 | 0.0000 | *** |
| | factor(location_id)10 | -14.300 | 2.794 | -5.119 | 0.0000 | *** |
| | factor(location_id)11 | -14.460 | 2.794 | -5.176 | 0.0000 | *** |
| | factor(location_id)12 | -9.500 | 2.794 | -3.401 | 0.0009 | *** |
| | factor(location_id)13 | -13.840 | 2.794 | -4.954 | 0.0000 | *** |
| | factor(location_id)14 | -5.060 | 2.794 | -1.811 | 0.0727 | . |
| | factor(location_id)15 | -4.940 | 2.794 | -1.768 | 0.0797 | . |
| | factor(location_id)16 | -11.880 | 2.794 | -4.253 | 0.0000 | *** |
| | factor(location_id)17 | -10.860 | 2.794 | -3.888 | 0.0002 | *** |
| | factor(location_id)18 | -8.280 | 2.794 | -2.964 | 0.0037 | ** |
| | factor(location_id)19 | -11.080 | 2.794 | -3.966 | 0.0001 | *** |
| | factor(location_id)20 | -9.780 | 2.794 | -3.501 | 0.0007 | *** |
| | factor(location_id)21 | -11.980 | 2.794 | -4.288 | 0.0000 | *** |
| | factor(location_id)22 | -7.880 | 2.794 | -2.821 | 0.0057 | ** |
| | factor(location_id)23 | -7.640 | 2.794 | -2.735 | 0.0072 | ** |
| | factor(location_id)24 | -9.720 | 2.794 | -3.479 | 0.0007 | *** |
| | factor(location_id)25 | -16.000 | 2.794 | -5.728 | 0.0000 | *** |
| | factor(location_id)26 | -5.840 | 2.794 | -2.091 | 0.0388 | * |
| | factor(location_id)27 | -9.560 | 2.794 | -3.422 | 0.0009 | *** |
| | factor(location_id)28 | -17.190 | 3.230 | -5.322 | 0.0000 | *** |
| | factor(location_id)29 | -13.520 | 2.794 | -4.840 | 0.0000 | *** |
| | factor(location_id)281 | -16.348 | 3.238 | -5.049 | 0.0000 | *** |
| | factor(location_id)282 | -18.775 | 4.849 | -3.872 | 0.0002 | *** |

| Component | Term | Estimate | Std Error | t-value | p-value | |
|---|---|---|---|---|---|---|
| **Component** | **Term** | **edf** | **Ref. df** | **F-value** | **p-value** | |
| B. smooth terms | s(n) | 1.214 | 1.388 | 18.485 | 0.0000 | *** |

Signif. codes: 0 <= '***' < 0.001 < '**' < 0.01 < '*' < 0.05

Adjusted R-squared: 0.450, Deviance explained 0.568

GCV : 25.033, Scale est: 19.510, N: 146

**Table S10.** Generalized Additive Model Summary for LAX Lden (dBA) noise levels with a smoothing spline on flight traffic (denoted as variable n).

| Component | Term | Estimate | Std Error | t-value | p-value | |
|---|---|---|---|---|---|---|
| A. parametric coefficients | (Intercept) | 78.095 | 0.353 | 221.334 | 0.0000 | *** |
| | factor(SiteID)ATH1 | -17.667 | 0.497 | -35.568 | 0.0000 | *** |
| | factor(SiteID)ATH2 | -12.221 | 0.446 | -27.373 | 0.0000 | *** |
| | factor(SiteID)DEL1 | -20.421 | 0.446 | -45.740 | 0.0000 | *** |
| | factor(SiteID)ESG1 | -13.621 | 0.446 | -30.509 | 0.0000 | *** |
| | factor(SiteID)ESG2 | -10.621 | 0.446 | -23.790 | 0.0000 | *** |
| | factor(SiteID)ESG3 | -16.021 | 0.446 | -35.885 | 0.0000 | *** |
| | factor(SiteID)ESG4 | -19.000 | 0.497 | -38.252 | 0.0000 | *** |
| | factor(SiteID)ESG5 | -17.421 | 0.446 | -39.020 | 0.0000 | *** |
| | factor(SiteID)ING1 | -17.421 | 0.446 | -39.020 | 0.0000 | *** |
| | factor(SiteID)ING2 | -12.421 | 0.446 | -27.821 | 0.0000 | *** |
| | factor(SiteID)ING3 | -11.821 | 0.446 | -26.477 | 0.0000 | *** |
| | factor(SiteID)ING5 | -18.667 | 0.497 | -37.581 | 0.0000 | *** |
| | factor(SiteID)ING6 | -8.621 | 0.446 | -19.310 | 0.0000 | *** |
| | factor(SiteID)ING7 | -21.333 | 0.497 | -42.950 | 0.0000 | *** |
| | factor(SiteID)ING8 | -16.821 | 0.446 | -37.676 | 0.0000 | *** |
| | factor(SiteID)LNX1 | -4.021 | 0.446 | -9.007 | 0.0000 | *** |
| | factor(SiteID)LNX2 | -15.621 | 0.446 | -34.989 | 0.0000 | *** |
| | factor(SiteID)LNX3 | -15.821 | 0.446 | -35.437 | 0.0000 | *** |
| | factor(SiteID)LNX4 | -13.021 | 0.446 | -29.165 | 0.0000 | *** |
| | factor(SiteID)PDR1 | -10.421 | 0.446 | -23.342 | 0.0000 | *** |
| | factor(SiteID)PDR2 | -16.021 | 0.446 | -35.885 | 0.0000 | *** |
| | factor(SiteID)PDR3 | -20.333 | 0.497 | -40.937 | 0.0000 | *** |
| | factor(SiteID)SLA1 | -13.821 | 0.446 | -30.957 | 0.0000 | *** |
| | factor(SiteID)SLA2 | -19.333 | 0.497 | -38.923 | 0.0000 | *** |
| | factor(SiteID)SLA3 | -16.421 | 0.446 | -36.780 | 0.0000 | *** |
| | factor(SiteID)SLA4 | -19.000 | 0.497 | -38.252 | 0.0000 | *** |
| | factor(SiteID)SLA5 | -14.021 | 0.446 | -31.405 | 0.0000 | *** |
| | factor(SiteID)SLA6 | -16.333 | 0.497 | -32.884 | 0.0000 | *** |
| | factor(SiteID)SLA7 | -13.821 | 0.446 | -30.957 | 0.0000 | *** |
| | factor(SiteID)SLA8 | -17.333 | 0.497 | -34.897 | 0.0000 | *** |

| Component | Term | Estimate | Std Error | t-value | p-value | |
|---|---|---|---|---|---|---|
| | factor(SiteID)SLA9 | -17.000 | 0.497 | -34.226 | 0.0000 | *** |
| | factor(SiteID)WCH1 | -21.333 | 0.497 | -42.950 | 0.0000 | *** |
| | factor(SiteID)WCH2 | -16.221 | 0.446 | -36.333 | 0.0000 | *** |
| | factor(SiteID)WCH3 | -17.021 | 0.446 | -38.124 | 0.0000 | *** |
| | factor(SiteID)WCH4 | -20.000 | 0.497 | -40.266 | 0.0000 | *** |
| | factor(SiteID)WCH5 | -5.021 | 0.446 | -11.247 | 0.0000 | *** |
| | factor(SiteID)WCH6 | -15.621 | 0.446 | -34.989 | 0.0000 | *** |
| **Component** | **Term** | **edf** | **Ref. df** | **F-value** | **p-value** | |
| B. smooth terms | s(n) | 3.045 | 3.244 | 186.056 | 0.0000 | *** |

Signif. codes: 0 <= '***' < 0.001 < '**' < 0.01 < '*' < 0.05

Adjusted R-squared: 0.983, Deviance explained 0.987

GCV : 0.494, Scale est: 0.370, N: 164

**Table S11.** Generalized Additive Model Summary for ORD Ldn (dBA) noise levels with a smoothing spline on flight traffic (denoted as variable n).

| Component | Term | Estimate | Std Error | t-value | p-value | |
|---|---|---|---|---|---|---|
| A. parametric coefficients | (Intercept) | 52.194 | 0.587 | 88.948 | 0.0000 | *** |
| | factor(SiteID)10 | -4.020 | 0.830 | -4.844 | 0.0000 | *** |
| | factor(SiteID)11 | 0.760 | 0.830 | 0.916 | 0.3614 | |
| | factor(SiteID)12 | 5.620 | 0.830 | 6.772 | 0.0000 | *** |
| | factor(SiteID)13 | 10.960 | 0.830 | 13.207 | 0.0000 | *** |
| | factor(SiteID)14 | 2.600 | 0.830 | 3.133 | 0.0021 | ** |
| | factor(SiteID)15 | 0.100 | 0.830 | 0.121 | 0.9043 | |
| | factor(SiteID)16 | 11.460 | 0.830 | 13.810 | 0.0000 | *** |
| | factor(SiteID)17 | 9.820 | 0.830 | 11.833 | 0.0000 | *** |
| | factor(SiteID)18 | 5.909 | 0.881 | 6.710 | 0.0000 | *** |
| | factor(SiteID)19 | 2.580 | 0.830 | 3.109 | 0.0023 | ** |
| | factor(SiteID)2 | 2.240 | 0.830 | 2.699 | 0.0078 | ** |
| | factor(SiteID)20 | 0.880 | 0.830 | 1.060 | 0.2908 | |
| | factor(SiteID)21 | -2.421 | 0.880 | -2.751 | 0.0068 | ** |
| | factor(SiteID)22 | 12.440 | 0.830 | 14.991 | 0.0000 | *** |
| | factor(SiteID)23 | 7.840 | 0.830 | 9.447 | 0.0000 | *** |
| | factor(SiteID)24 | 1.820 | 0.830 | 2.193 | 0.0300 | * |
| | factor(SiteID)25 | 9.980 | 0.830 | 12.026 | 0.0000 | *** |
| | factor(SiteID)26 | 4.680 | 0.830 | 5.640 | 0.0000 | *** |
| | factor(SiteID)27 | 16.000 | 0.830 | 19.280 | 0.0000 | *** |
| | factor(SiteID)28 | 20.880 | 0.830 | 25.161 | 0.0000 | *** |
| | factor(SiteID)29 | 14.760 | 0.830 | 17.786 | 0.0000 | *** |
| | factor(SiteID)3 | 11.380 | 0.830 | 13.713 | 0.0000 | *** |
| | factor(SiteID)30 | 3.579 | 0.880 | 4.066 | 0.0001 | *** |
| | factor(SiteID)31 | 18.434 | 0.881 | 20.933 | 0.0000 | *** |
| | factor(SiteID)32 | 11.500 | 0.830 | 13.858 | 0.0000 | *** |
| | factor(SiteID)33 | 9.020 | 0.830 | 10.869 | 0.0000 | *** |
| | factor(SiteID)34 | 5.240 | 0.830 | 6.314 | 0.0000 | *** |
| | factor(SiteID)35 | 3.740 | 0.830 | 4.507 | 0.0000 | *** |
| | factor(SiteID)37 | -3.480 | 0.830 | -4.194 | 0.0000 | *** |
| | factor(SiteID)39 | 8.186 | 1.438 | 5.694 | 0.0000 | *** |

| Component | Term | Estimate | Std Error | t-value | p-value | |
|---|---|---|---|---|---|---|
| | factor(SiteID)4 | 3.420 | 0.830 | 4.121 | 0.0001 | *** |
| | factor(SiteID)40 | 2.186 | 1.438 | 1.521 | 0.1307 | |
| | factor(SiteID)42 | 4.860 | 0.830 | 5.856 | 0.0000 | *** |
| | factor(SiteID)44 | 11.209 | 0.881 | 12.729 | 0.0000 | *** |
| | factor(SiteID)45 | -1.114 | 1.438 | -0.775 | 0.4396 | |
| | factor(SiteID)46 | 3.986 | 1.438 | 2.773 | 0.0063 | ** |
| | factor(SiteID)5 | 5.679 | 0.880 | 6.451 | 0.0000 | *** |
| | factor(SiteID)7 | -0.846 | 0.880 | -0.962 | 0.3379 | |
| | factor(SiteID)8 | 5.000 | 0.830 | 6.025 | 0.0000 | *** |
| **Component** | **Term** | **edf** | **Ref. df** | **F-value** | **p-value** | |
| B. smooth terms | s(n) | 1.000 | 1.000 | 208.612 | 0.0000 | *** |

Signif. codes: 0 <= '***' < 0.001 < '**' < 0.01 < '*' < 0.05

Adjusted R-squared: 0.955, Deviance explained 0.965

GCV : 2.241, Scale est: 1.722, N: 177

**Table S12.** Generalized Additive Model Summary for JFK, LGA, and EWR Ldn (dBA) noise levels with a smoothing spline on flight traffic (denoted as variable n).

| Component | Term | Estimate | Std Error | t-value | p-value | |
|---|---|---|---|---|---|---|
| A. parametric coefficients | (Intercept) | 70.160 | 0.629 | 111.529 | 0.0000 | *** |
| | factor(SiteID)2 | 0.080 | 0.890 | 0.090 | 0.9285 | |
| | factor(SiteID)3 | -25.100 | 0.890 | -28.213 | 0.0000 | *** |
| | factor(SiteID)4 | -5.440 | 0.890 | -6.115 | 0.0000 | *** |
| | factor(SiteID)5 | 0.160 | 0.890 | 0.180 | 0.8576 | |
| | factor(SiteID)6 | -1.800 | 0.890 | -2.023 | 0.0452 | * |
| | factor(SiteID)7 | -8.260 | 0.890 | -9.285 | 0.0000 | *** |
| | factor(SiteID)8 | -5.160 | 0.890 | -5.800 | 0.0000 | *** |
| | factor(SiteID)9 | -3.700 | 0.890 | -4.159 | 0.0001 | *** |
| | factor(SiteID)11 | -1.980 | 0.890 | -2.226 | 0.0278 | * |
| | factor(SiteID)28 | -3.240 | 0.890 | -3.642 | 0.0004 | *** |
| | factor(SiteID)52 | -15.020 | 0.890 | -16.883 | 0.0000 | *** |
| | factor(SiteID)54 | -15.540 | 0.890 | -17.468 | 0.0000 | *** |
| | factor(SiteID)55 | -9.220 | 0.890 | -10.364 | 0.0000 | *** |
| | factor(SiteID)56 | -3.200 | 0.890 | -3.597 | 0.0005 | *** |
| | factor(SiteID)57 | -4.065 | 0.944 | -4.307 | 0.0000 | *** |
| | factor(SiteID)58 | -7.080 | 0.890 | -7.958 | 0.0000 | *** |
| | factor(SiteID)59 | -7.100 | 0.890 | -7.981 | 0.0000 | *** |
| | factor(SiteID)62 | -9.920 | 0.890 | -11.150 | 0.0000 | *** |
| | factor(SiteID)66 | -14.080 | 0.890 | -15.827 | 0.0000 | *** |
| | factor(SiteID)67 | -10.760 | 0.890 | -12.095 | 0.0000 | *** |
| | factor(SiteID)68 | -9.557 | 1.180 | -8.096 | 0.0000 | *** |
| | factor(SiteID)69 | -19.300 | 0.890 | -21.694 | 0.0000 | *** |
| | factor(SiteID)71 | -10.300 | 0.890 | -11.578 | 0.0000 | *** |
| | factor(SiteID)72 | -16.900 | 0.890 | -18.996 | 0.0000 | *** |
| | factor(SiteID)74 | -2.860 | 0.890 | -3.215 | 0.0017 | ** |
| | factor(SiteID)76 | -16.057 | 1.180 | -13.602 | 0.0000 | *** |
| | factor(SiteID)77 | -16.920 | 0.890 | -19.019 | 0.0000 | *** |
| | factor(SiteID)78 | -20.700 | 0.890 | -23.268 | 0.0000 | *** |
| | factor(SiteID)79 | -12.140 | 0.944 | -12.862 | 0.0000 | *** |
| | factor(SiteID)80 | -17.720 | 0.890 | -19.918 | 0.0000 | *** |

| Component | Term | Estimate | Std Error | t-value | p-value | |
|---|---|---|---|---|---|---|
| | factor(SiteID)82 | -3.200 | 0.890 | -3.597 | 0.0005 | *** |
| | factor(SiteID)84 | -16.328 | 0.944 | -17.298 | 0.0000 | *** |
| | factor(SiteID)85 | -19.224 | 1.029 | -18.680 | 0.0000 | *** |
| | factor(SiteID)86 | -15.239 | 1.543 | -9.874 | 0.0000 | *** |
| **Component** | **Term** | **edf** | **Ref. df** | **F-value** | **p-value** | |
| B. smooth terms | s(n) | 1.000 | 1.000 | 248.766 | 0.0000 | *** |

Signif. codes: 0 <= '***' < 0.001 < '**' < 0.01 < '*' < 0.05

Adjusted R-squared: 0.962, Deviance explained 0.971

GCV : 2.553, Scale est: 1.979, N: 160
