## Supplementary material for "Fewer flights and lower community noise: Noise monitoring at six airports in the United States before, during, and after the COVID-19 pandemic": LM results for year

**Table S5.** Linear Model Summary for SFO Lden (dBA) noise levels with categorical year.

| | Estimate | Standard Error | t value | Pr(>ltl) | |
|---|---|---|---|---|---|
| (Intercept) | 75.329 | 1.779 | 42.355 | 0.0000 | *** |
| factor(year)2018 | 0.279 | 1.202 | 0.232 | 0.8170 | |
| factor(year)2019 | -0.497 | 1.193 | -0.416 | 0.6777 | |
| factor(year)2020 | -4.876 | 1.187 | -4.109 | 0.0001 | *** |
| factor(year)2021 | -3.512 | 1.199 | -2.928 | 0.0038 | ** |
| factor(year)2022 | -3.352 | 1.193 | -2.810 | 0.0055 | ** |
| factor(year)2023 | -2.410 | 1.200 | -2.009 | 0.0459 | * |
| factor(year)2024 | -2.865 | 1.200 | -2.389 | 0.0179 | * |
| factor(location_id)2 | -8.527 | 2.435 | -3.502 | 0.0006 | *** |
| factor(location_id)3 | -10.050 | 2.248 | -4.470 | 0.0000 | *** |
| factor(location_id)4 | -5.650 | 2.248 | -2.513 | 0.0128 | * |
| factor(location_id)5 | -6.100 | 2.248 | -2.713 | 0.0073 | ** |
| factor(location_id)6 | -8.600 | 2.248 | -3.825 | 0.0002 | *** |
| factor(location_id)7 | -12.763 | 2.248 | -5.676 | 0.0000 | *** |
| factor(location_id)8 | -4.363 | 2.248 | -1.940 | 0.0538 | . |
| factor(location_id)9 | -14.362 | 2.248 | -6.388 | 0.0000 | *** |
| factor(location_id)10 | -14.225 | 2.248 | -6.326 | 0.0000 | *** |
| factor(location_id)11 | -14.550 | 2.248 | -6.471 | 0.0000 | *** |
| factor(location_id)12 | -9.737 | 2.248 | -4.331 | 0.0000 | *** |
| factor(location_id)13 | -14.863 | 2.248 | -6.610 | 0.0000 | *** |
| factor(location_id)14 | -6.888 | 2.248 | -3.063 | 0.0025 | ** |
| factor(location_id)15 | -7.337 | 2.248 | -3.263 | 0.0013 | ** |
| factor(location_id)16 | -8.213 | 2.248 | -3.652 | 0.0003 | *** |
| factor(location_id)17 | -11.212 | 2.248 | -4.987 | 0.0000 | *** |
| factor(location_id)18 | -7.850 | 2.248 | -3.491 | 0.0006 | *** |
| factor(location_id)19 | -11.113 | 2.248 | -4.942 | 0.0000 | *** |
| factor(location_id)20 | -10.525 | 2.248 | -4.681 | 0.0000 | *** |
| factor(location_id)21 | -12.338 | 2.248 | -5.487 | 0.0000 | *** |
| factor(location_id)22 | -8.175 | 2.248 | -3.636 | 0.0004 | *** |
| factor(location_id)23 | -8.238 | 2.248 | -3.664 | 0.0003 | *** |
| factor(location_id)24 | -9.112 | 2.248 | -4.053 | 0.0001 | *** |
| factor(location_id)25 | -15.988 | 2.248 | -7.110 | 0.0000 | *** |
| factor(location_id)26 | -8.025 | 2.248 | -3.569 | 0.0005 | *** |
| factor(location_id)27 | -11.250 | 2.248 | -5.003 | 0.0000 | *** |

| | Estimate | Standard Error | t value | Pr(>ltl) | |
|---|---|---|---|---|---|
| factor(location_id)28 | -10.406 | 2.770 | -3.756 | 0.0002 | *** |
| factor(location_id)29 | -13.888 | 2.248 | -6.176 | 0.0000 | *** |
| factor(location_id)281 | -16.549 | 3.069 | -5.393 | 0.0000 | *** |
| factor(location_id)282 | -18.920 | 3.069 | -6.165 | 0.0000 | *** |

*Signif. codes: 0 <= '***' < 0.001 < '**' < 0.01 < '*' < 0.05*

Residual standard error: 4.497 on 194 degrees of freedom

Multiple R-squared: 0.5055, Adjusted R-squared: 0.4112

F-statistic: 5.36 on 194 and 37 DF, p-value: 0.0000

**Table S6.** Linear Model Summary for LAX Lden (dBA) noise levels with categorical year.

| | Estimate | Standard Error | t value | Pr(>ltl) | |
|---|---|---|---|---|---|
| (Intercept) | 79.412 | 0.360 | 220.528 | 0.0000 | *** |
| factor(year)2019 | -0.263 | 0.139 | -1.887 | 0.0616 | . |
| factor(year)2020 | -2.974 | 0.139 | -21.322 | 0.0000 | *** |
| factor(year)2021 | -1.919 | 0.162 | -11.857 | 0.0000 | *** |
| factor(year)2022 | -1.799 | 0.162 | -11.115 | 0.0000 | *** |
| factor(SiteID)ATH1 | -17.667 | 0.496 | -35.592 | 0.0000 | *** |
| factor(SiteID)ATH2 | -12.221 | 0.446 | -27.391 | 0.0000 | *** |
| factor(SiteID)DEL1 | -20.421 | 0.446 | -45.770 | 0.0000 | *** |
| factor(SiteID)ESG1 | -13.621 | 0.446 | -30.529 | 0.0000 | *** |
| factor(SiteID)ESG2 | -10.621 | 0.446 | -23.805 | 0.0000 | *** |
| factor(SiteID)ESG3 | -16.021 | 0.446 | -35.908 | 0.0000 | *** |
| factor(SiteID)ESG4 | -19.000 | 0.496 | -38.278 | 0.0000 | *** |
| factor(SiteID)ESG5 | -17.421 | 0.446 | -39.046 | 0.0000 | *** |
| factor(SiteID)ING1 | -17.421 | 0.446 | -39.046 | 0.0000 | *** |
| factor(SiteID)ING2 | -12.421 | 0.446 | -27.839 | 0.0000 | *** |
| factor(SiteID)ING3 | -11.821 | 0.446 | -26.495 | 0.0000 | *** |
| factor(SiteID)ING5 | -18.667 | 0.496 | -37.607 | 0.0000 | *** |
| factor(SiteID)ING6 | -8.621 | 0.446 | -19.323 | 0.0000 | *** |
| factor(SiteID)ING7 | -21.333 | 0.496 | -42.979 | 0.0000 | *** |
| factor(SiteID)ING8 | -16.821 | 0.446 | -37.701 | 0.0000 | *** |
| factor(SiteID)LNX1 | -4.021 | 0.446 | -9.013 | 0.0000 | *** |
| factor(SiteID)LNX2 | -15.621 | 0.446 | -35.012 | 0.0000 | *** |
| factor(SiteID)LNX3 | -15.821 | 0.446 | -35.460 | 0.0000 | *** |
| factor(SiteID)LNX4 | -13.021 | 0.446 | -29.184 | 0.0000 | *** |
| factor(SiteID)PDR1 | -10.421 | 0.446 | -23.357 | 0.0000 | *** |
| factor(SiteID)PDR2 | -16.021 | 0.446 | -35.908 | 0.0000 | *** |
| factor(SiteID)PDR3 | -20.333 | 0.496 | -40.964 | 0.0000 | *** |
| factor(SiteID)SLA1 | -13.821 | 0.446 | -30.977 | 0.0000 | *** |
| factor(SiteID)SLA2 | -19.333 | 0.496 | -38.950 | 0.0000 | *** |
| factor(SiteID)SLA3 | -16.421 | 0.446 | -36.805 | 0.0000 | *** |
| factor(SiteID)SLA4 | -19.000 | 0.496 | -38.278 | 0.0000 | *** |
| factor(SiteID)SLA5 | -14.021 | 0.446 | -31.425 | 0.0000 | *** |
| factor(SiteID)SLA6 | -16.333 | 0.496 | -32.906 | 0.0000 | *** |
| factor(SiteID)SLA7 | -13.821 | 0.446 | -30.977 | 0.0000 | *** |

| | Estimate | Standard Error | t value | Pr(>|t|) |
|---|---|---|---|---|---|
| factor(SiteID)SLA8 | -17.333 | 0.496 | -34.921 | 0.0000 | *** |
| factor(SiteID)SLA9 | -17.000 | 0.496 | -34.249 | 0.0000 | *** |
| factor(SiteID)WCH1 | -21.333 | 0.496 | -42.979 | 0.0000 | *** |
| factor(SiteID)WCH2 | -16.221 | 0.446 | -36.356 | 0.0000 | *** |
| factor(SiteID)WCH3 | -17.021 | 0.446 | -38.149 | 0.0000 | *** |
| factor(SiteID)WCH4 | -20.000 | 0.496 | -40.293 | 0.0000 | *** |
| factor(SiteID)WCH5 | -5.021 | 0.446 | -11.254 | 0.0000 | *** |
| factor(SiteID)WCH6 | -15.621 | 0.446 | -35.012 | 0.0000 | *** |

*Signif. codes: 0 <= '***' < 0.001 < '**' < 0.01 < '*' < 0.05*

Residual standard error: 0.6079 on 122 degrees of freedom

Multiple R-squared: 0.987, Adjusted R-squared: 0.9826

F-statistic: 225.4 on 122 and 41 DF, p-value: 0.0000

**Table S7.** Linear Model Summary for ORD Ldn (dBA) noise levels with categorical year.

| | Estimate | Standard Error | t value | Pr(>|t|) |
|---|---|---|---|---|---|
| (Intercept) | 54.104 | 0.615 | 88.022 | 0.0000 | *** |
| factor(year)2019 | -0.543 | 0.314 | -1.727 | 0.0866 | . |
| factor(year)2020 | -4.059 | 0.314 | -12.913 | 0.0000 | *** |
| factor(year)2021 | -2.515 | 0.314 | -8.000 | 0.0000 | *** |
| factor(year)2022 | -2.305 | 0.326 | -7.063 | 0.0000 | *** |
| factor(SiteID)10 | -4.020 | 0.821 | -4.899 | 0.0000 | *** |
| factor(SiteID)11 | 0.760 | 0.821 | 0.926 | 0.3560 | |
| factor(SiteID)12 | 5.620 | 0.821 | 6.849 | 0.0000 | *** |
| factor(SiteID)13 | 10.960 | 0.821 | 13.357 | 0.0000 | *** |
| factor(SiteID)14 | 2.600 | 0.821 | 3.169 | 0.0019 | ** |
| factor(SiteID)15 | 0.100 | 0.821 | 0.122 | 0.9032 | |
| factor(SiteID)16 | 11.460 | 0.821 | 13.966 | 0.0000 | *** |
| factor(SiteID)17 | 9.820 | 0.821 | 11.967 | 0.0000 | *** |
| factor(SiteID)18 | 6.001 | 0.872 | 6.883 | 0.0000 | *** |
| factor(SiteID)19 | 2.580 | 0.821 | 3.144 | 0.0021 | ** |
| factor(SiteID)2 | 2.240 | 0.821 | 2.730 | 0.0072 | ** |
| factor(SiteID)20 | 0.880 | 0.821 | 1.072 | 0.2855 | |
| factor(SiteID)21 | -2.450 | 0.872 | -2.810 | 0.0057 | ** |
| factor(SiteID)22 | 12.440 | 0.821 | 15.160 | 0.0000 | *** |
| factor(SiteID)23 | 7.840 | 0.821 | 9.554 | 0.0000 | *** |
| factor(SiteID)24 | 1.820 | 0.821 | 2.218 | 0.0283 | * |
| factor(SiteID)25 | 9.980 | 0.821 | 12.162 | 0.0000 | *** |
| factor(SiteID)26 | 4.680 | 0.821 | 5.703 | 0.0000 | *** |
| factor(SiteID)27 | 16.000 | 0.821 | 19.499 | 0.0000 | *** |
| factor(SiteID)28 | 20.880 | 0.821 | 25.446 | 0.0000 | *** |
| factor(SiteID)29 | 14.760 | 0.821 | 17.988 | 0.0000 | *** |
| factor(SiteID)3 | 11.380 | 0.821 | 13.869 | 0.0000 | *** |
| factor(SiteID)30 | 3.550 | 0.872 | 4.072 | 0.0001 | *** |
| factor(SiteID)31 | 18.526 | 0.872 | 21.250 | 0.0000 | *** |
| factor(SiteID)32 | 11.500 | 0.821 | 14.015 | 0.0000 | *** |
| factor(SiteID)33 | 9.020 | 0.821 | 10.992 | 0.0000 | *** |
| factor(SiteID)34 | 5.240 | 0.821 | 6.386 | 0.0000 | *** |
| factor(SiteID)35 | 3.740 | 0.821 | 4.558 | 0.0000 | *** |
| factor(SiteID)37 | -3.480 | 0.821 | -4.241 | 0.0000 | *** |

| | Estimate | Standard Error | t value | Pr(>ltl) | |
|---|---|---|---|---|---|
| factor(SiteID)39 | 8.300 | 1.436 | 5.780 | 0.0000 | *** |
| factor(SiteID)4 | 3.420 | 0.821 | 4.168 | 0.0001 | *** |
| factor(SiteID)40 | 2.300 | 1.436 | 1.602 | 0.1116 | |
| factor(SiteID)42 | 4.860 | 0.821 | 5.923 | 0.0000 | *** |
| factor(SiteID)44 | 11.301 | 0.872 | 12.963 | 0.0000 | *** |
| factor(SiteID)45 | -1.000 | 1.436 | -0.696 | 0.4875 | |
| factor(SiteID)46 | 4.100 | 1.436 | 2.855 | 0.0050 | ** |
| factor(SiteID)5 | 5.650 | 0.872 | 6.480 | 0.0000 | *** |
| factor(SiteID)7 | -0.875 | 0.872 | -1.004 | 0.3174 | |
| factor(SiteID)8 | 5.000 | 0.821 | 6.093 | 0.0000 | *** |

*Signif. codes: 0 <= '***' < 0.001 < '**' < 0.01 < '*' < 0.05*

Residual standard error: 1.297 on 133 degrees of freedom

Multiple R-squared: 0.9668, Adjusted R-squared: 0.9561

F-statistic: 90.06 on 133 and 43 DF, p-value: 0.0000

**Table S8.** Linear Model Summary for JFK, LGA, and EWR Ldn (dBA) noise levels with categorical year.

| | Estimate | Standard Error | t value | Pr(>ltl) | |
|---|---|---|---|---|---|
| (Intercept) | 72.248 | 0.607 | 119.107 | 0.0000 | *** |
| factor(year)2019 | -0.271 | 0.316 | -0.858 | 0.3925 | |
| factor(year)2020 | -4.748 | 0.323 | -14.689 | 0.0000 | *** |
| factor(year)2021 | -3.645 | 0.323 | -11.277 | 0.0000 | *** |
| factor(year)2022 | -1.773 | 0.332 | -5.342 | 0.0000 | *** |
| factor(SiteID)2 | 0.080 | 0.807 | 0.099 | 0.9212 | |
| factor(SiteID)3 | -25.100 | 0.807 | -31.086 | 0.0000 | *** |
| factor(SiteID)4 | -5.440 | 0.807 | -6.737 | 0.0000 | *** |
| factor(SiteID)5 | 0.160 | 0.807 | 0.198 | 0.8433 | |
| factor(SiteID)6 | -1.800 | 0.807 | -2.229 | 0.0276 | * |
| factor(SiteID)7 | -8.260 | 0.807 | -10.230 | 0.0000 | *** |
| factor(SiteID)8 | -5.160 | 0.807 | -6.391 | 0.0000 | *** |
| factor(SiteID)9 | -3.700 | 0.807 | -4.582 | 0.0000 | *** |
| factor(SiteID)11 | -1.980 | 0.807 | -2.452 | 0.0156 | * |
| factor(SiteID)28 | -3.240 | 0.807 | -4.013 | 0.0001 | *** |
| factor(SiteID)52 | -15.020 | 0.807 | -18.602 | 0.0000 | *** |
| factor(SiteID)54 | -15.540 | 0.807 | -19.246 | 0.0000 | *** |
| factor(SiteID)55 | -9.220 | 0.807 | -11.419 | 0.0000 | *** |
| factor(SiteID)56 | -3.200 | 0.807 | -3.963 | 0.0001 | *** |
| factor(SiteID)57 | -4.331 | 0.858 | -5.048 | 0.0000 | *** |
| factor(SiteID)58 | -7.080 | 0.807 | -8.768 | 0.0000 | *** |
| factor(SiteID)59 | -7.100 | 0.807 | -8.793 | 0.0000 | *** |
| factor(SiteID)62 | -9.920 | 0.807 | -12.286 | 0.0000 | *** |
| factor(SiteID)66 | -14.080 | 0.807 | -17.438 | 0.0000 | *** |
| factor(SiteID)67 | -10.760 | 0.807 | -13.326 | 0.0000 | *** |
| factor(SiteID)68 | -10.062 | 1.076 | -9.355 | 0.0000 | *** |
| factor(SiteID)69 | -19.300 | 0.807 | -23.903 | 0.0000 | *** |
| factor(SiteID)71 | -10.300 | 0.807 | -12.756 | 0.0000 | *** |
| factor(SiteID)72 | -16.900 | 0.807 | -20.930 | 0.0000 | *** |
| factor(SiteID)74 | -2.860 | 0.807 | -3.542 | 0.0006 | *** |
| factor(SiteID)76 | -16.562 | 1.076 | -15.399 | 0.0000 | *** |
| factor(SiteID)77 | -16.920 | 0.807 | -20.955 | 0.0000 | *** |
| factor(SiteID)78 | -20.700 | 0.807 | -25.637 | 0.0000 | *** |
| factor(SiteID)79 | -12.406 | 0.858 | -14.459 | 0.0000 | *** |
| factor(SiteID)80 | -17.720 | 0.807 | -21.946 | 0.0000 | *** |
| factor(SiteID)82 | -3.200 | 0.807 | -3.963 | 0.0001 | *** |
| factor(SiteID)84 | -16.188 | 0.858 | -18.868 | 0.0000 | *** |
| factor(SiteID)85 | -19.393 | 0.936 | -20.716 | 0.0000 | *** |
| factor(SiteID)86 | -14.174 | 1.415 | -10.020 | 0.0000 | *** |

*Signif. codes: 0 <= '***' < 0.001 < '**' < 0.01 < '*' < 0.05*

Residual standard error: 1.277 on 121 degrees of freedom

Multiple R-squared: 0.9764, Adjusted R-squared: 0.9689

F-statistic: 131.6 on 121 and 38 DF, p-value: 0.0000
