## Supplementary material for "Fewer flights and lower community noise: Noise monitoring at six airports in the United States before, during, and after the COVID-19 pandemic": LM results for restricted model without 2020 flight traffic

**Table S18.** Linear Model Summary for SFO Lden (dBA) noise levels with continuous flight traffic (denoted as per 1000 flights, variable nper1000), restricted model with year 2020 removed.

| | Estimate | Standard Error | t value | Pr(>ltl) | |
|---|---|---|---|---|---|
| (Intercept) | 65.266 | 3.294 | 19.815 | 0.0000 | *** |
| nper1000 | 0.027 | 0.007 | 3.612 | 0.0005 | *** |
| factor(location_id)2 | -8.474 | 3.808 | -2.225 | 0.0287 | * |
| factor(location_id)3 | -10.025 | 3.522 | -2.846 | 0.0056 | ** |
| factor(location_id)4 | -5.600 | 3.522 | -1.590 | 0.1156 | |
| factor(location_id)5 | -5.975 | 3.522 | -1.696 | 0.0935 | . |
| factor(location_id)6 | -8.475 | 3.522 | -2.406 | 0.0183 | * |
| factor(location_id)7 | -11.675 | 3.522 | -3.314 | 0.0014 | ** |
| factor(location_id)8 | -2.600 | 3.522 | -0.738 | 0.4625 | |
| factor(location_id)9 | -14.150 | 3.522 | -4.017 | 0.0001 | *** |
| factor(location_id)10 | -14.250 | 3.522 | -4.046 | 0.0001 | *** |
| factor(location_id)11 | -14.475 | 3.522 | -4.109 | 0.0001 | *** |
| factor(location_id)12 | -9.400 | 3.522 | -2.669 | 0.0091 | ** |
| factor(location_id)13 | -13.400 | 3.522 | -3.804 | 0.0003 | *** |
| factor(location_id)14 | -3.925 | 3.522 | -1.114 | 0.2683 | |
| factor(location_id)15 | -3.375 | 3.522 | -0.958 | 0.3407 | |
| factor(location_id)16 | -11.800 | 3.522 | -3.350 | 0.0012 | ** |
| factor(location_id)17 | -10.600 | 3.522 | -3.009 | 0.0035 | ** |
| factor(location_id)18 | -8.225 | 3.522 | -2.335 | 0.0219 | * |
| factor(location_id)19 | -10.950 | 3.522 | -3.109 | 0.0026 | ** |
| factor(location_id)20 | -9.325 | 3.522 | -2.647 | 0.0097 | ** |
| factor(location_id)21 | -11.875 | 3.522 | -3.371 | 0.0011 | ** |
| factor(location_id)22 | -8.000 | 3.522 | -2.271 | 0.0257 | * |
| factor(location_id)23 | -7.300 | 3.522 | -2.072 | 0.0413 | * |
| factor(location_id)24 | -9.425 | 3.522 | -2.676 | 0.0090 | ** |
| factor(location_id)25 | -15.925 | 3.522 | -4.521 | 0.0000 | *** |
| factor(location_id)26 | -4.275 | 3.522 | -1.214 | 0.2283 | |

| | Estimate | Standard Error | t value | Pr(>|t|) |
|---|---|---|---|---|---|
| factor(location_id)27 | -8.350 | 3.522 | -2.371 | 0.0201 | * |
| factor(location_id)28 | -18.196 | 4.337 | -4.196 | 0.0001 | *** |
| factor(location_id)29 | -13.675 | 3.522 | -3.882 | 0.0002 | *** |
| factor(location_id)281 | -16.754 | 4.337 | -3.863 | 0.0002 | *** |
| factor(location_id)282 | -18.697 | 5.573 | -3.355 | 0.0012 | ** |

*Signif. codes: 0 <= '***' < 0.001 < '**' < 0.01 < '*' < 0.05*

Residual standard error: 4.981 on 84 degrees of freedom

Multiple R-squared: 0.5372, Adjusted R-squared: 0.3664

F-statistic: 3.145 on 84 and 31 DF, p-value: 0.0000

**Table S19.** Linear Model Summary for LAX Lden (dBA) noise levels with continuous flight traffic (denoted as per 1000 flights, variable nper1000), restricted model with year 2020 removed.

| | Estimate | Standard Error | t value | Pr(>ltl) | |
|---|---|---|---|---|---|
| (Intercept) | 69.723 | 0.842 | 82.819 | 0.0000 | *** |
| nper1000 | 0.021 | 0.002 | 13.067 | 0.0000 | *** |
| factor(SiteID)ATH1 | -17.500 | 0.640 | -27.353 | 0.0000 | *** |
| factor(SiteID)ATH2 | -11.953 | 0.557 | -21.443 | 0.0000 | *** |
| factor(SiteID)DEL1 | -20.453 | 0.557 | -36.692 | 0.0000 | *** |
| factor(SiteID)ESG1 | -13.453 | 0.557 | -24.134 | 0.0000 | *** |
| factor(SiteID)ESG2 | -10.453 | 0.557 | -18.752 | 0.0000 | *** |
| factor(SiteID)ESG3 | -15.953 | 0.557 | -28.619 | 0.0000 | *** |
| factor(SiteID)ESG4 | -19.000 | 0.640 | -29.697 | 0.0000 | *** |
| factor(SiteID)ESG5 | -17.453 | 0.557 | -31.310 | 0.0000 | *** |
| factor(SiteID)ING1 | -17.203 | 0.557 | -30.862 | 0.0000 | *** |
| factor(SiteID)ING2 | -11.953 | 0.557 | -21.443 | 0.0000 | *** |
| factor(SiteID)ING3 | -11.453 | 0.557 | -20.546 | 0.0000 | *** |
| factor(SiteID)ING5 | -18.500 | 0.640 | -28.916 | 0.0000 | *** |
| factor(SiteID)ING6 | -8.453 | 0.557 | -15.164 | 0.0000 | *** |
| factor(SiteID)ING7 | -20.500 | 0.640 | -32.042 | 0.0000 | *** |
| factor(SiteID)ING8 | -16.703 | 0.557 | -29.965 | 0.0000 | *** |
| factor(SiteID)LNX1 | -3.953 | 0.557 | -7.091 | 0.0000 | *** |
| factor(SiteID)LNX2 | -15.453 | 0.557 | -27.722 | 0.0000 | *** |
| factor(SiteID)LNX3 | -15.703 | 0.557 | -28.171 | 0.0000 | *** |
| factor(SiteID)LNX4 | -12.953 | 0.557 | -23.237 | 0.0000 | *** |
| factor(SiteID)PDR1 | -9.953 | 0.557 | -17.855 | 0.0000 | *** |
| factor(SiteID)PDR2 | -15.703 | 0.557 | -28.171 | 0.0000 | *** |
| factor(SiteID)PDR3 | -20.000 | 0.640 | -31.260 | 0.0000 | *** |
| factor(SiteID)SLA1 | -13.453 | 0.557 | -24.134 | 0.0000 | *** |
| factor(SiteID)SLA2 | -19.000 | 0.640 | -29.697 | 0.0000 | *** |
| factor(SiteID)SLA3 | -16.203 | 0.557 | -29.068 | 0.0000 | *** |

| | Estimate | Standard Error | t value | Pr(>ltl) | |
|---|---|---|---|---|---|
| factor(SiteID)SLA4 | -18.500 | 0.640 | -28.916 | 0.0000 | *** |
| factor(SiteID)SLA5 | -13.953 | 0.557 | -25.031 | 0.0000 | *** |
| factor(SiteID)SLA6 | -16.000 | 0.640 | -25.008 | 0.0000 | *** |
| factor(SiteID)SLA7 | -13.703 | 0.557 | -24.583 | 0.0000 | *** |
| factor(SiteID)SLA8 | -17.500 | 0.640 | -27.353 | 0.0000 | *** |
| factor(SiteID)SLA9 | -16.500 | 0.640 | -25.790 | 0.0000 | *** |
| factor(SiteID)WCH1 | -21.000 | 0.640 | -32.823 | 0.0000 | *** |
| factor(SiteID)WCH2 | -15.953 | 0.557 | -28.619 | 0.0000 | *** |
| factor(SiteID)WCH3 | -16.703 | 0.557 | -29.965 | 0.0000 | *** |
| factor(SiteID)WCH4 | -19.500 | 0.640 | -30.479 | 0.0000 | *** |
| factor(SiteID)WCH5 | -4.703 | 0.557 | -8.437 | 0.0000 | *** |
| factor(SiteID)WCH6 | -15.203 | 0.557 | -27.274 | 0.0000 | *** |

*Signif. codes: 0 <= '***' < 0.001 < '**' < 0.01 < '*' < 0.05*

Residual standard error: 0.6398 on 87 degrees of freedom

Multiple R-squared: 0.9852, Adjusted R-squared: 0.9787

F-statistic: 152 on 87 and 38 DF, p-value: 0.0000

**Table S20.** Linear Model Summary for ORD Ldn (dBA) noise levels with continuous flight traffic (denoted as per 1000 flights, variable nper1000), restricted model with year 2020 removed.

| | Estimate | Standard Error | t value | Pr(>ltl) | |
|---|---|---|---|---|---|
| (Intercept) | 45.636 | 0.959 | 47.572 | 0.0000 | *** |
| nper1000 | 0.012 | 0.001 | 9.677 | 0.0000 | *** |
| factor(SiteID)10 | -4.025 | 0.889 | -4.530 | 0.0000 | *** |
| factor(SiteID)11 | 0.600 | 0.889 | 0.675 | 0.5011 | |
| factor(SiteID)12 | 6.000 | 0.889 | 6.752 | 0.0000 | *** |
| factor(SiteID)13 | 11.375 | 0.889 | 12.801 | 0.0000 | *** |
| factor(SiteID)14 | 2.475 | 0.889 | 2.785 | 0.0064 | ** |
| factor(SiteID)15 | 0.125 | 0.889 | 0.141 | 0.8884 | |
| factor(SiteID)16 | 11.225 | 0.889 | 12.632 | 0.0000 | *** |
| factor(SiteID)17 | 9.775 | 0.889 | 11.000 | 0.0000 | *** |
| factor(SiteID)18 | 5.597 | 0.960 | 5.828 | 0.0000 | *** |
| factor(SiteID)19 | 2.725 | 0.889 | 3.067 | 0.0028 | ** |
| factor(SiteID)2 | 2.875 | 0.889 | 3.235 | 0.0016 | ** |
| factor(SiteID)20 | 1.000 | 0.889 | 1.125 | 0.2631 | |
| factor(SiteID)21 | -2.279 | 0.960 | -2.373 | 0.0195 | * |
| factor(SiteID)22 | 12.350 | 0.889 | 13.898 | 0.0000 | *** |
| factor(SiteID)23 | 7.900 | 0.889 | 8.890 | 0.0000 | *** |
| factor(SiteID)24 | 2.125 | 0.889 | 2.391 | 0.0187 | * |
| factor(SiteID)25 | 10.600 | 0.889 | 11.929 | 0.0000 | *** |
| factor(SiteID)26 | 4.900 | 0.889 | 5.514 | 0.0000 | *** |
| factor(SiteID)27 | 17.100 | 0.889 | 19.243 | 0.0000 | *** |
| factor(SiteID)28 | 20.775 | 0.889 | 23.379 | 0.0000 | *** |
| factor(SiteID)29 | 14.775 | 0.889 | 16.627 | 0.0000 | *** |
| factor(SiteID)3 | 11.275 | 0.889 | 12.688 | 0.0000 | *** |
| factor(SiteID)30 | 3.454 | 0.960 | 3.598 | 0.0005 | *** |
| factor(SiteID)31 | 18.531 | 0.960 | 19.296 | 0.0000 | *** |
| factor(SiteID)32 | 11.625 | 0.889 | 13.082 | 0.0000 | *** |

| | Estimate | Standard Error | t value | Pr(>ltl) | |
|---|---|---|---|---|---|
| factor(SiteID)33 | 9.750 | 0.889 | 10.972 | 0.0000 | *** |
| factor(SiteID)34 | 5.000 | 0.889 | 5.627 | 0.0000 | *** |
| factor(SiteID)35 | 3.850 | 0.889 | 4.333 | 0.0000 | *** |
| factor(SiteID)37 | -4.175 | 0.889 | -4.698 | 0.0000 | *** |
| factor(SiteID)39 | 8.237 | 1.407 | 5.853 | 0.0000 | *** |
| factor(SiteID)4 | 3.575 | 0.889 | 4.023 | 0.0001 | *** |
| factor(SiteID)40 | 2.237 | 1.407 | 1.590 | 0.1151 | |
| factor(SiteID)42 | 5.150 | 0.889 | 5.796 | 0.0000 | *** |
| factor(SiteID)44 | 11.097 | 0.960 | 11.555 | 0.0000 | *** |
| factor(SiteID)45 | -1.063 | 1.407 | -0.755 | 0.4519 | |
| factor(SiteID)46 | 4.037 | 1.407 | 2.868 | 0.0050 | ** |
| factor(SiteID)5 | 6.388 | 0.960 | 6.652 | 0.0000 | *** |
| factor(SiteID)7 | -0.946 | 0.960 | -0.985 | 0.3271 | |
| factor(SiteID)8 | 5.025 | 0.889 | 5.655 | 0.0000 | *** |

*Signif. codes: 0 <= '***' < 0.001 < '**' < 0.01 < '*' < 0.05*

Residual standard error: 1.257 on 100 degrees of freedom

Multiple R-squared: 0.9703, Adjusted R-squared: 0.9585

F-statistic: 81.81 on 100 and 40 DF, p-value: 0.0000

**Table S21.** Linear Model Summary for JFK, LGA, and EWR Ldn (dBA) noise levels with continuous flight traffic (denoted as per 1000 flights, variable nper1000), restricted model with year 2020 removed.

| | Estimate | Standard Error | t value | Pr(>ltl) | |
|---|---|---|---|---|---|
| (Intercept) | 64.020 | 1.011 | 63.327 | 0.0000 | *** |
| nper1000 | 0.008 | 0.001 | 9.810 | 0.0000 | *** |
| factor(SiteID)11 | -1.675 | 1.057 | -1.585 | 0.1164 | |
| factor(SiteID)2 | -0.125 | 1.057 | -0.118 | 0.9061 | |
| factor(SiteID)28 | -3.275 | 1.057 | -3.099 | 0.0026 | ** |
| factor(SiteID)3 | -24.700 | 1.057 | -23.376 | 0.0000 | *** |
| factor(SiteID)4 | -5.450 | 1.057 | -5.158 | 0.0000 | *** |
| factor(SiteID)5 | 0.175 | 1.057 | 0.166 | 0.8688 | |
| factor(SiteID)52 | -14.875 | 1.057 | -14.078 | 0.0000 | *** |
| factor(SiteID)54 | -15.275 | 1.057 | -14.456 | 0.0000 | *** |
| factor(SiteID)55 | -9.325 | 1.057 | -8.825 | 0.0000 | *** |
| factor(SiteID)56 | -3.100 | 1.057 | -2.934 | 0.0042 | ** |
| factor(SiteID)57 | -3.863 | 1.142 | -3.384 | 0.0011 | ** |
| factor(SiteID)58 | -6.525 | 1.057 | -6.175 | 0.0000 | *** |
| factor(SiteID)59 | -6.975 | 1.057 | -6.601 | 0.0000 | *** |
| factor(SiteID)6 | -1.700 | 1.057 | -1.609 | 0.1111 | |
| factor(SiteID)62 | -9.700 | 1.057 | -9.180 | 0.0000 | *** |
| factor(SiteID)66 | -14.025 | 1.057 | -13.273 | 0.0000 | *** |
| factor(SiteID)67 | -10.775 | 1.057 | -10.197 | 0.0000 | *** |
| factor(SiteID)68 | -9.428 | 1.297 | -7.272 | 0.0000 | *** |
| factor(SiteID)69 | -19.000 | 1.057 | -17.981 | 0.0000 | *** |
| factor(SiteID)7 | -8.500 | 1.057 | -8.044 | 0.0000 | *** |
| factor(SiteID)71 | -10.200 | 1.057 | -9.653 | 0.0000 | *** |
| factor(SiteID)72 | -17.000 | 1.057 | -16.089 | 0.0000 | *** |
| factor(SiteID)74 | -2.825 | 1.057 | -2.674 | 0.0089 | ** |
| factor(SiteID)76 | -15.928 | 1.297 | -12.285 | 0.0000 | *** |
| factor(SiteID)77 | -16.400 | 1.057 | -15.521 | 0.0000 | *** |

| | Estimate | Standard Error | t value | Pr(>ltl) | |
|---|---|---|---|---|---|
| factor(SiteID)78 | -19.650 | 1.057 | -18.597 | 0.0000 | *** |
| factor(SiteID)79 | -11.863 | 1.142 | -10.392 | 0.0000 | *** |
| factor(SiteID)8 | -5.250 | 1.057 | -4.969 | 0.0000 | *** |
| factor(SiteID)80 | -17.800 | 1.057 | -16.846 | 0.0000 | *** |
| factor(SiteID)82 | -3.100 | 1.057 | -2.934 | 0.0042 | ** |
| factor(SiteID)84 | -16.013 | 1.142 | -14.026 | 0.0000 | *** |
| factor(SiteID)85 | -19.415 | 1.297 | -14.974 | 0.0000 | *** |
| factor(SiteID)86 | -15.110 | 1.672 | -9.036 | 0.0000 | *** |
| factor(SiteID)9 | -3.725 | 1.057 | -3.525 | 0.0007 | *** |

*Signif. codes: 0 <= '***' < 0.001 < '**' < 0.01 < '*' < 0.05*

Residual standard error: 1.494 on 92 degrees of freedom

Multiple R-squared: 0.9673, Adjusted R-squared: 0.9549

F-statistic: 77.77 on 92 and 35 DF, p-value: 0.0000
