## Supplementary material for "Fewer flights and lower community noise: Noise monitoring at six airports in the United States before, during, and after the COVID-19 pandemic": LM results for flight traffic

**Table S13.** Linear Model Summary for SFO Lden (dBA) noise levels with continuous flight traffic (denoted as per 1000 flights, variable nper1000).

| | Estimate | Standard Error | t value | Pr(>ltl) | |
|---|---|---|---|---|---|
| (Intercept) | 65.499 | 2.412 | 27.152 | 0.0000 | *** |
| nper1000 | 0.027 | 0.005 | 5.213 | 0.0000 | *** |
| factor(location_id)2 | -8.470 | 2.969 | -2.853 | 0.0051 | ** |
| factor(location_id)3 | -9.640 | 2.797 | -3.447 | 0.0008 | *** |
| factor(location_id)4 | -5.720 | 2.797 | -2.045 | 0.0431 | * |
| factor(location_id)5 | -6.000 | 2.797 | -2.145 | 0.0341 | * |
| factor(location_id)6 | -8.600 | 2.797 | -3.075 | 0.0026 | ** |
| factor(location_id)7 | -11.900 | 2.797 | -4.255 | 0.0000 | *** |
| factor(location_id)8 | -2.900 | 2.797 | -1.037 | 0.3020 | |
| factor(location_id)9 | -13.960 | 2.797 | -4.991 | 0.0000 | *** |
| factor(location_id)10 | -14.300 | 2.797 | -5.113 | 0.0000 | *** |
| factor(location_id)11 | -14.460 | 2.797 | -5.170 | 0.0000 | *** |
| factor(location_id)12 | -9.500 | 2.797 | -3.397 | 0.0009 | *** |
| factor(location_id)13 | -13.840 | 2.797 | -4.948 | 0.0000 | *** |
| factor(location_id)14 | -5.060 | 2.797 | -1.809 | 0.0731 | . |
| factor(location_id)15 | -4.940 | 2.797 | -1.766 | 0.0800 | . |
| factor(location_id)16 | -11.880 | 2.797 | -4.248 | 0.0000 | *** |
| factor(location_id)17 | -10.860 | 2.797 | -3.883 | 0.0002 | *** |
| factor(location_id)18 | -8.280 | 2.797 | -2.961 | 0.0037 | ** |
| factor(location_id)19 | -11.080 | 2.797 | -3.962 | 0.0001 | *** |
| factor(location_id)20 | -9.780 | 2.797 | -3.497 | 0.0007 | *** |
| factor(location_id)21 | -11.980 | 2.797 | -4.283 | 0.0000 | *** |
| factor(location_id)22 | -7.880 | 2.797 | -2.817 | 0.0057 | ** |
| factor(location_id)23 | -7.640 | 2.797 | -2.732 | 0.0073 | ** |
| factor(location_id)24 | -9.720 | 2.797 | -3.475 | 0.0007 | *** |
| factor(location_id)25 | -16.000 | 2.797 | -5.721 | 0.0000 | *** |
| factor(location_id)26 | -5.840 | 2.797 | -2.088 | 0.0390 | * |

| | Estimate | Standard Error | t value | Pr(>ltl) | |
|---|---|---|---|---|---|
| factor(location_id)27 | -9.560 | 2.797 | -3.418 | 0.0009 | *** |
| factor(location_id)28 | -17.139 | 3.232 | -5.303 | 0.0000 | *** |
| factor(location_id)29 | -13.520 | 2.797 | -4.834 | 0.0000 | *** |
| factor(location_id)281 | -16.375 | 3.242 | -5.052 | 0.0000 | *** |
| factor(location_id)282 | -18.936 | 4.844 | -3.909 | 0.0002 | *** |

*Signif. codes: 0 <= '***' < 0.001 < '**' < 0.01 < '*' < 0.05*

Residual standard error: 4.422 on 114 degrees of freedom

Multiple R-squared: 0.5664, Adjusted R-squared: 0.4484

F-statistic: 4.803 on 114 and 31 DF, p-value: 0.0000

**Table S14.** Linear Model Summary for LAX Lden (dBA) noise levels with continuous flight traffic (denoted as per 1000 flights, variable nper1000).

| | Estimate | Standard Error | t value | Pr(>ltl) | |
|---|---|---|---|---|---|
| (Intercept) | 72.755 | 0.477 | 152.671 | 0.0000 | *** |
| nper1000 | 0.015 | 0.001 | 21.118 | 0.0000 | *** |
| factor(SiteID)ATH1 | -17.667 | 0.561 | -31.496 | 0.0000 | *** |
| factor(SiteID)ATH2 | -12.465 | 0.502 | -24.845 | 0.0000 | *** |
| factor(SiteID)DEL1 | -20.665 | 0.502 | -41.190 | 0.0000 | *** |
| factor(SiteID)ESG1 | -13.865 | 0.502 | -27.636 | 0.0000 | *** |
| factor(SiteID)ESG2 | -10.865 | 0.502 | -21.656 | 0.0000 | *** |
| factor(SiteID)ESG3 | -16.265 | 0.502 | -32.420 | 0.0000 | *** |
| factor(SiteID)ESG4 | -19.000 | 0.561 | -33.873 | 0.0000 | *** |
| factor(SiteID)ESG5 | -17.665 | 0.502 | -35.210 | 0.0000 | *** |
| factor(SiteID)ING1 | -17.665 | 0.502 | -35.210 | 0.0000 | *** |
| factor(SiteID)ING2 | -12.665 | 0.502 | -25.244 | 0.0000 | *** |
| factor(SiteID)ING3 | -12.065 | 0.502 | -24.048 | 0.0000 | *** |
| factor(SiteID)ING5 | -18.667 | 0.561 | -33.279 | 0.0000 | *** |
| factor(SiteID)ING6 | -8.865 | 0.502 | -17.670 | 0.0000 | *** |
| factor(SiteID)ING7 | -21.333 | 0.561 | -38.033 | 0.0000 | *** |
| factor(SiteID)ING8 | -17.065 | 0.502 | -34.014 | 0.0000 | *** |
| factor(SiteID)LNX1 | -4.265 | 0.502 | -8.501 | 0.0000 | *** |
| factor(SiteID)LNX2 | -15.865 | 0.502 | -31.622 | 0.0000 | *** |
| factor(SiteID)LNX3 | -16.065 | 0.502 | -32.021 | 0.0000 | *** |
| factor(SiteID)LNX4 | -13.265 | 0.502 | -26.440 | 0.0000 | *** |
| factor(SiteID)PDR1 | -10.665 | 0.502 | -21.258 | 0.0000 | *** |
| factor(SiteID)PDR2 | -16.265 | 0.502 | -32.420 | 0.0000 | *** |
| factor(SiteID)PDR3 | -20.333 | 0.561 | -36.250 | 0.0000 | *** |
| factor(SiteID)SLA1 | -14.065 | 0.502 | -28.035 | 0.0000 | *** |
| factor(SiteID)SLA2 | -19.333 | 0.561 | -34.468 | 0.0000 | *** |
| factor(SiteID)SLA3 | -16.665 | 0.502 | -33.217 | 0.0000 | *** |

| | Estimate | Standard Error | t value | Pr(>ltl) | |
|---|---|---|---|---|---|
| factor(SiteID)SLA4 | -19.000 | 0.561 | -33.873 | 0.0000 | *** |
| factor(SiteID)SLA5 | -14.265 | 0.502 | -28.433 | 0.0000 | *** |
| factor(SiteID)SLA6 | -16.333 | 0.561 | -29.119 | 0.0000 | *** |
| factor(SiteID)SLA7 | -14.065 | 0.502 | -28.035 | 0.0000 | *** |
| factor(SiteID)SLA8 | -17.333 | 0.561 | -30.902 | 0.0000 | *** |
| factor(SiteID)SLA9 | -17.000 | 0.561 | -30.308 | 0.0000 | *** |
| factor(SiteID)WCH1 | -21.333 | 0.561 | -38.033 | 0.0000 | *** |
| factor(SiteID)WCH2 | -16.465 | 0.502 | -32.818 | 0.0000 | *** |
| factor(SiteID)WCH3 | -17.265 | 0.502 | -34.413 | 0.0000 | *** |
| factor(SiteID)WCH4 | -20.000 | 0.561 | -35.656 | 0.0000 | *** |
| factor(SiteID)WCH5 | -5.265 | 0.502 | -10.494 | 0.0000 | *** |
| factor(SiteID)WCH6 | -15.865 | 0.502 | -31.622 | 0.0000 | *** |

*Signif. codes: 0 <= '***' < 0.001 < '**' < 0.01 < '*' < 0.05*

Residual standard error: 0.687 on 125 degrees of freedom

Multiple R-squared: 0.983, Adjusted R-squared: 0.9778

F-statistic: 189.7 on 125 and 38 DF, p-value: 0.0000

**Table S15.** Linear Model Summary for ORD Ldn (dBA) noise levels with continuous flight traffic (denoted as per 1000 flights, variable nper1000).

*Signif. codes: 0 <= '***' < 0.001 < '**' < 0.01 < '*' < 0.05*

Residual standard error: 1.312 on 136 degrees of freedom

Multiple R-squared: 0.9653, Adjusted R-squared: 0.9551

F-statistic: 94.51 on 136 and 40 DF, p-value: 0.0000

**Table S16.** Linear Model Summary for JFK, LGA, and EWR Ldn (dBA) noise levels with continuous flight traffic (denoted as per 1000 flights, variable nper1000).

*Signif. codes: 0 <= '***' < 0.001 < '**' < 0.01 < '*' < 0.05*

Residual standard error: 1.407 on 124 degrees of freedom

Multiple R-squared: 0.9706, Adjusted R-squared: 0.9623

F-statistic: 117 on 124 and 35 DF, p-value: 0.0000
