## Supplementary material for "Fewer flights and lower community noise: Noise monitoring at six airports in the United States before, during, and after the COVID-19 pandemic": LM results for restricted model for SFO

**Table S22.** Linear Model Summary for SFO Lden (dBA) noise levels with continuous flight traffic (denoted as per 1000 flights, variable nper1000), restricted model with year 2020 removed, and 3 high dB sites removed.

| | Estimate | Standard Error | t value | Pr(>ltl) | |
|---|---|---|---|---|---|
| (Intercept) | 68.560 | 1.789 | 38.331 | 0.0000 | *** |
| nper1000 | 0.016 | 0.004 | 3.716 | 0.0004 | *** |
| factor(location_id)2 | -8.718 | 2.017 | -4.323 | 0.0000 | *** |
| factor(location_id)3 | -10.025 | 1.865 | -5.375 | 0.0000 | *** |
| factor(location_id)4 | -5.600 | 1.865 | -3.002 | 0.0036 | ** |
| factor(location_id)5 | -5.975 | 1.865 | -3.204 | 0.0020 | ** |
| factor(location_id)6 | -8.475 | 1.865 | -4.544 | 0.0000 | *** |
| factor(location_id)7 | -11.675 | 1.865 | -6.260 | 0.0000 | *** |
| factor(location_id)8 | -2.600 | 1.865 | -1.394 | 0.1674 | |
| factor(location_id)9 | -14.150 | 1.865 | -7.587 | 0.0000 | *** |
| factor(location_id)10 | -14.250 | 1.865 | -7.640 | 0.0000 | *** |
| factor(location_id)11 | -14.475 | 1.865 | -7.761 | 0.0000 | *** |
| factor(location_id)12 | -9.400 | 1.865 | -5.040 | 0.0000 | *** |
| factor(location_id)13 | -13.400 | 1.865 | -7.185 | 0.0000 | *** |
| factor(location_id)16 | -11.800 | 1.865 | -6.327 | 0.0000 | *** |
| factor(location_id)17 | -10.600 | 1.865 | -5.683 | 0.0000 | *** |
| factor(location_id)18 | -8.225 | 1.865 | -4.410 | 0.0000 | *** |
| factor(location_id)19 | -10.950 | 1.865 | -5.871 | 0.0000 | *** |
| factor(location_id)20 | -9.325 | 1.865 | -5.000 | 0.0000 | *** |
| factor(location_id)21 | -11.875 | 1.865 | -6.367 | 0.0000 | *** |
| factor(location_id)22 | -8.000 | 1.865 | -4.289 | 0.0001 | *** |
| factor(location_id)23 | -7.300 | 1.865 | -3.914 | 0.0002 | *** |
| factor(location_id)24 | -9.425 | 1.865 | -5.053 | 0.0000 | *** |
| factor(location_id)25 | -15.925 | 1.865 | -8.538 | 0.0000 | *** |
| factor(location_id)27 | -8.350 | 1.865 | -4.477 | 0.0000 | *** |
| factor(location_id)28 | -17.521 | 2.298 | -7.625 | 0.0000 | *** |
| factor(location_id)29 | -13.675 | 1.865 | -7.332 | 0.0000 | *** |
| factor(location_id)281 | -17.429 | 2.298 | -7.586 | 0.0000 | *** |
| factor(location_id)282 | -19.016 | 2.951 | -6.443 | 0.0000 | *** |

| | Estimate | Standard Error | t value | Pr(>|t|) |
|---|---|---|---|---|

_Signif. codes: 0 <= '***' < 0.001 < '**' < 0.01 < '*' < 0.05_

Residual standard error: 2.638 on 75 degrees of freedom

Multiple R-squared: 0.7714, Adjusted R-squared: 0.686

F-statistic: 9.037 on 75 and 28 DF, p-value: 0.0000
