## Supplementary material for "Fewer flights and lower community noise: Noise monitoring at six airports in the United States before, during, and after the COVID-19 pandemic": GAM and LM model comparisons

**Table S17.** Comparison of linear and generalized additive models with year and flight explanatory variables.

| Airports | Models | AIC | $R^2$ |
|---|---|---|---|
| SFO | LM year | 1,392.47 | 0.51 |
| | GAM year | 1,391.92 | 0.41 |
| | LM flights | 878.30 | 0.57 |
| | GAM flights | 878.11 | 0.45 |
| LAX | LM year | 339.64 | 0.99 |
| | GAM year | 462.19 | 0.96 |
| | LM flights | 377.73 | 0.98 |
| | GAM flights | 339.24 | 0.98 |
| ORD | LM year | 633.89 | 0.97 |
| | GAM year | 695.36 | 0.94 |
| | LM flights | 635.83 | 0.97 |
| | GAM flights | 635.83 | 0.96 |
| JFK, LGA, EWR | LM year | 567.52 | 0.98 |
| | GAM year | 622.96 | 0.96 |
| | LM flights | 596.47 | 0.97 |
| | GAM flights | 596.47 | 0.96 |
